# Mono and biallelic variants in *HCN2* cause severe neurodevelopmental disorders

**DOI:** 10.1101/2024.03.19.24303984

**Authors:** Clara Houdayer, A Marie Phillips, Marie Chabbert, Jennifer Bourreau, Reza Maroofian, Henry Houlden, Kay Richards, Nebal Waill Saadi, Eliška Dad’ová, Patrick Van Bogaert, Mailys Rupin, Boris Keren, Perrine Charles, Thomas Smol, Audrey Riquet, Lynn Pais, Anne O’Donnell-Luria, Grace E. VanNoy, Allan Bayat, Rikke S Møller, Kern Olofsson, Rami Abou Jamra, Steffen Syrbe, Majed Dasouki, Laurie H Seaver, Jennifer A Sullivan, Vandana Shashi, Fowzan S Alkuraya, Alexis F Poss, J Edward Spence, Rhonda E Schnur, Ian C Forster, Chaseley E Mckenzie, Cas Simons, Min Wang, Penny Snell, Kavitha Kothur, Michael Buckley, Tony Roscioli, Noha Elserafy, Benjamin Dauriat, Vincent Procaccio, Daniel Henrion, Guy Lenaers, Estelle Colin, Nienke E. Verbeek, Koen L. Van Gassen, Claire Legendre, Dominique Bonneau, Christopher A Reid, Katherine B Howell, Alban Ziegler, Christian Legros

## Abstract

Hyperpolarization activated Cyclic Nucleotide (HCN) gated channels are crucial for various neurophysiological functions, including learning and sensory functions, and their dysfunction are responsible for brain disorders, such as epilepsy. To date, *HCN2* variants have only been associated with mild epilepsy and recently, one monoallelic missense variant has been linked to developmental and epileptic encephalopathy. Here, we expand the phenotypic spectrum of *HCN2-*related disorders by describing twenty-one additional individuals from fifteen unrelated families carrying *HCN2* variants. Seventeen individuals had developmental delay/intellectual disability (DD/ID), two had borderline DD/ID, and one had borderline DD. Ten individuals had epilepsy with DD/ID, with median age of onset of 10 months, and one had epilepsy with normal development. Molecular diagnosis identified thirteen different pathogenic *HCN2* variants, including eleven missense variants affecting highly conserved amino acids, one frameshift variant, and one in-frame deletion. Seven variants were monoallelic of which five occurred *de novo,* one was not maternally inherited, one was inherited from a father with mild learning disabilities, and one was of unknown inheritance. The remaining six variants were biallelic, with four homozygous and two compound heterozygous variants. Functional studies using two-electrode voltage-clamp recordings in *Xenopus laevis* oocytes were performed on three monoallelic variants, p.(Arg324His), p.(Ala363Val), and p.(Met374Leu), and three biallelic variants, p.(Leu377His), p.(Pro493Leu) and p.(Gly587Asp). The p.(Arg324His) variant induced a strong increase of HCN2 conductance, while p.(Ala363Val) and p.(Met374Leu) displayed dominant negative effects, leading to a partial loss of HCN2 channel function. By confocal imaging, we found that the p.(Leu377His), p.(Pro493Leu) and p.(Gly587Asp) pathogenic variants impaired membrane trafficking, resulting in a complete loss of HCN2 elicited currents in *Xenopus* oocytes. Structural 3D-analysis in depolarized and hyperpolarized states of HCN2 channels, revealed that the pathogenic variants p.(His205Gln), p.(Ser409Leu), p.(Arg324Cys), p.(Asn369Ser) and p.(Gly460Asp) modify molecular interactions altering HCN2 function. Taken together, our data broadens the clinical spectrum associated with *HCN2* variants, and disclose that *HCN2* is involved in developmental encephalopathy with or without epilepsy.

## Introduction

Diseases resulting from dysfunction of ion channels, collectively known as channelopathies, often manifest as neurodevelopmental disorders associated with epilepsy.^1–3^ Among the ion channels involved in epilepsy, the Hyperpolarization activated Cyclic Nucleotide (HCN) gated ion channels, have been highlighted as important regulators of neuronal excitability in disease for more than two decades.^4–7^ The human genome contains four distinct HCN genes (*HCN1-4*) that have a high level of sequence homology, but exhibit different cerebral expression patterns. *HCN1* and *HCN2* are the most abundant and widely expressed isoforms in brain, while *HCN3* is predominant in the cerebellum, and *HCN4* is weakly expressed in the brain but highly abundant in the heart.^4,8^

HCN 1-4 channels are composed of four subunits forming a central ion-conducting pore and assembled in homo- or heterotetramers.^9^ Their structure-function relationships have been extensively investigated,^10^ and the cryo-EM structure of the human HCN1 channel has provided insight into the gating and ion selectivity mechanisms at the molecular level.^11^ HCN1-4 channels open upon hyperpolarization of the membrane to generate a Na^+^ inward current, called I_h_ and I_f_ in neurons^12,13^ and cardiomyocytes^14,15^ respectively, leading to membrane depolarization in excitable cells and exhibiting pacemaker activity. These ion channels are potentiated by intracellular cyclic monophosphate nucleotides cAMP and cGMP, through a positive shift of activation range.^4,16^ The I_h_ current is involved in resting potential regulation, dendritic integration, and synaptic transmission.^4^ Several studies performed on transgenic mice have established a relationship between *HCN1* and *HCN2* and neuronal excitability and epileptogenesis, and being implicated in various neurophysiological processes, including sleep, learning, memory, and sensory functions.^17–20^

Of the HCN channels, HCN1 has been extensively investigated in human neurological disease. Heterozygous missense *HCN1* variants were first reported in individuals with developmental and epileptic encephalopathy (DEE) (DEE24; # 615871).^21–23^ Thereafter, *HCN1* has been associated with a broader clinical spectrum including febrile seizures, milder epilepsies (including genetic generalized epilepsies (GGE) and genetic epilepsy with febrile seizures plus (GEFS+)), and intellectual disability (ID) without seizures.^23^ A genotype/phenotype correlation has been identified, with *HCN1* variants located in the transmembrane domain being associated with DEE, while those located in the N- or C-terminal mostly resulted in milder epilepsy.^23^

To date, six pathogenic *HCN2* variants have been reported in a restricted number of individuals, with mild epilepsies including GGE and GEFS+, mostly without ID (Table 1, Fig. 1A). ^24–27^ Only two individuals with *HCN2* variants had ID: one of the three individuals from the family carrying the p.(Val246Met) variant, and one girl presenting ID and epilepsy, caused by a *de novo* p.(Gly460Asp) variant.^27,28^ Electrophysiological studies have revealed that most *HCN2* variants studied to date p.(Pro719-Pro721del) (previously reported as delPPP), p.(Ser632Trp) and p.(Val246Met) produce a gain-of-function (GOF) of the HCN2 channel, due to either larger current densities, faster kinetics, or a positive shift of voltage-dependent activation.^27,29^ The biallelic p.(Glu515Lys) and the monoallelic p.(Gly460Asp) variants have been reported to cause loss-of-function (LOF); the first, by slowing activation kinetics and shifting activation to more hyperpolarized potentials, resulting in an absence of current^25^ and the second, by inducing a strong reduction of the current density secondary due to trafficking defect to the cell membrane.^28^

**Figure 1.**
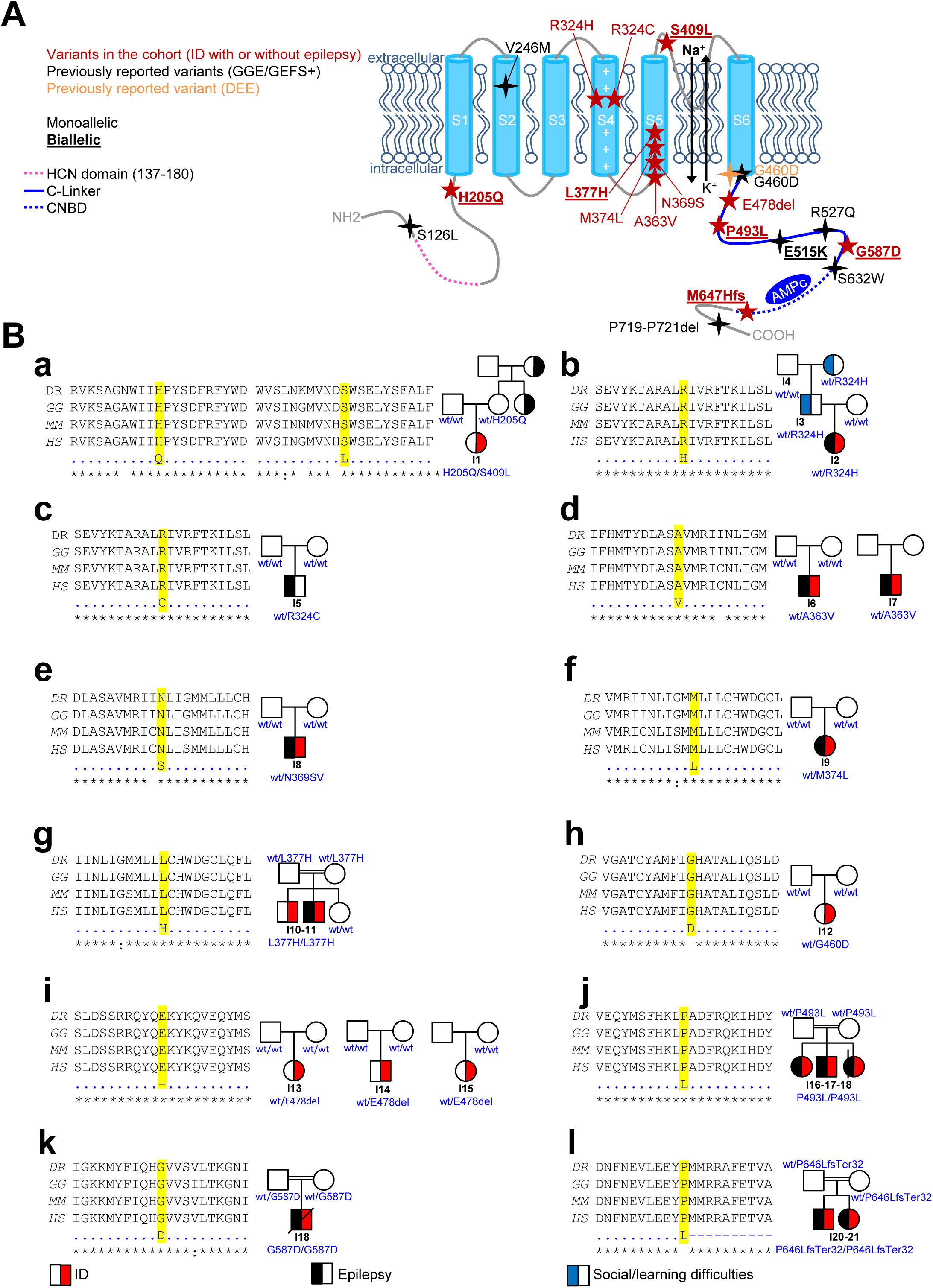
Pedigrees and *HCN2* pathogenic variants. **(A)** HCN2 channel and pathogenic variants. Variants identified in the cohort are indicated by red stars, variants previously reported are indicated by black stars. Biallelic variants are underlined while monoallelic are not. Pink dotted line, green full line and green dotted line indicate HCN domain, C-linker, and CNBD, respectively. **(B)** Pedigrees of I1–21 and multi-species sequence alignment of the mutated HCN2 channels. **(a-i)** Pedigree and multi-species sequence alignment of I1-21. Multi-species sequence alignment was performed using Clustal Omega {Uniprot entries: A0A8N7T6V3 (*Danio rerio*), A0A8V1AGR5 (*Gallus gallus*), O88703 (*Mus musculus*), Q9UL51 (*Homo sapiens*)}. Below the sequence alignment, a key denotes conserved amino acid (*), conservative replacement (:) and non-conservative replacement (). When not mentioned, the individual’s genotype is unknown.

In this study, we report twenty-one individuals from fifteen unrelated families, carrying twelve novel mono- or biallelic *HCN2* variants. By characterizing the biophysical properties of six variants by two-electrode voltage-clamp method (TEVC), Western blot analysis and 3D modeling, we demonstrate that *HCN2* variants are associated with a more severe and broader clinical spectrum of neurodevelopmental disorders than previously reported, and confirm its involvement in DEE.

## Materials and methods

### Individual recruitment and clinical phenotyping

This study was performed in accordance with ethical principles for medical research outlined in the Declaration of Helsinki. Individuals harboring *HCN2* pathogenic or likely pathogenic variants were recruited through an international collaborative effort facilitated by GeneMatcher^30^. Consents for publication of clinical data including genetic analyses were obtained from all individuals or their parents.

Individuals were enrolled from epilepsy and genetic centers located in Australia, Denmark, France, Germany, Netherlands, Saudi Arabia, UK, and USA. Physicians recorded clinical, EEG, neuroimaging and genetic data using a dedicated form. Epilepsy syndromes and seizure type were classified according to the International League Against Epilepsy (ILAE) guidelines.^31^

### Genetic analysis

Genetic analyses were performed in a diagnostic or research laboratory in all individuals, and when available, in their parents. *HCN2* variants were identified using either proband-only or Trio-exome or genome sequencing and confirmed by Sanger sequencing. Variants were classified according to ACMG criteria.^32^ The consequences of *HCN2* variants were interpreted on isoform NM_001194.4 (Supplementary Table 1).

#### TEVC recording of wt-HCN 2 and its variants

##### *HCN2* mutagenesis and *in vitro* transcription

A synthetic wild-type HCN2 cDNA (Genecust, Boynes, France) corresponding to the coding wild-type sequence of *HCN2* (wt-HCN2) (NM_001194.4) was inserted between the *BamH*I and *Hind*III sites in pGEM-HEJUEL, an expression vector suitable for *Xenopus laevis* oocyte expression. The *HCN2* variants, p.(Arg324His), p.(Ala363Val), p.(Met374Leu), p.(Leu377His), p.(Pro493Leu) and p.(Gly587Asp), were generated by site-mutagenesis, and their sequences were verified by Sanger sequencing. mRNAs were prepared from linearized cDNA templates by *in vitro* transcription using the mMESSAGE mMACHINE® T7 transcription Kit (Ambion, Fisher Scientific S.A.S., Illkirch, France) and purified with the NucleoSpin RNA clean up kits (Macherey Nagel, Düren, Germany) or RNeasyPlus (Qiagen, Germany) and quantified by nanospectrophotometry and stored at −24°C or −80°C until used.

##### Oocyte preparation and mRNA injection

All animal procedures were performed in accordance with the European Community council directive 2010/63/EU for the care and use of laboratory animals, and approved by the French Ministry of Agriculture (authorization N°A49007002 and APAFIS N#19433-2019022511329240v2). The NC3R’s ARRIVE guidelines were followed in the conduct of all experiments using animals. Adult *Xenopus laevis* females were used for oocytes collection as previously described. ^33,34^ For HCN2 expression, 21 ng of mRNA (wt-HCN2 or its variants) were injected into individual defolliculated oocytes, using an automatic nanoinjector (Nanoject II Drummond Scientific) or the Roboinject (Multi Channel Systems, Reutlingen, Germany). To mimic heterozygosity, heterotetrameric channels (e.g. wt/p.(Arg324His)) were produced by co-expression experiments with equal amounts (10.5 ng) of wt-*HCN2* and *HCN2* variant mRNAs. Injected oocytes were incubated 3-4 days at 18°C prior TEVC recordings.

##### Two-Electrode Voltage Clamp Recording

Details of solutions used for TEVC recordings can be found in the Supplementary material. Non-injected oocytes were used as negative control. From a holding potential, Vh = −30 mV, currents were elicited with a two-step protocol, comprising: i) an activation step in 10 mV increments from −140 mV to −30 mV for 8 s and ii) a return to −130 mV for 3 s. The first step of this protocol was used to establish the current-voltage relationships after current densities calculation with membrane capacitance. The second step was used to characterize the voltage-dependency of activation after normalization with maximum current amplitude. Currents were analysed using Clampfit 10.7 (Molecular Devices).

##### Western blot analysis

Details of solutions and Western blotting procedures are available in the Supplementary material. Thirty µg of protein from control and experimental samples (equivalent to protein from 80%-100% of protein from one oocyte) were separated by electrophoresis on 8% polyacrylamide SDS-PAGE gels. Proteins were transferred to nitrocellulose and blots were blocked with a milk-containing solution before incubation with 1:500 rabbit anti-HCN2 APC-030 (Alomone Labs Cat# APC-030, RRID: AB2313726). Beta-actin (Thermo Fisher Scientific Cat# PA1-183, RRID:AB_2539914) was used as a loading control. The protein signal was visualized with Clarity Western ECL Substrate (Bio-Rad, CA, USA) and the signal captured by a Bio-Rad ChemiDoc™ MP imaging system (Image Lab Software, RRID: SCR_014210).

##### Transient expression in HEK293 cells and confocal fluorescent microscopy

Details of solutions and procedure are available in the Supplementary material. HEK293T cells were transfected with N-terminal EGFP tagged constructs encoding human wt-HCN2, p.(Leu377His), p.(Pro493Leu) and p.(Gly587Asp). As negative control, a truncated HCN2_ΔC-X_ (Thr553Ter), a trafficking-deficient construct, was used.^35^ The membrane was labelled with CellMask Orange Plasma Membrane. The nuclei were stained with DAPI. Images were collected at 20x overview and then at high resolution (magnification objective: 63x) using Airy scan. The imager was “blind” to the genotype of the constructs.

##### Protein 3D modelling and structural analysis

Molecular models of depolarized and hyperpolarized wild type HCN2 were built with MODELLER V9.17 from position 163, by homology with the cryo-electron microscopy of human HCN1 (PDB 5U6P and 6UQF corresponding to depolarized and hyperpolarized models, respectively) (Supplementary Fig. 1).^11,36^ These templates were selected after comparison with rabbit HCN4 (PDB 7NMN).^37^ HCN1 and HCN4 structures differ in the S4-S5 linker, with an Mg2+ binding site observed only in HCN4.^38^ For each modelling, twenty structures were built and refined by simulated annealing (refine.fast option). The final models were selected according to the MODELLER PDFf score. The wt-HCN2 3D-models were subsequently used as templates for modeling the p.(His205Gln), p.(Arg324His), p.(Arg324Cys), p.(Ala363Val), p.(Asn369Ser), p.(Met374Leu), p.(Leu377His), p.(Gly460Asp), p.(Glu478del), p.(Pro493Leu) and p.(Gly587Asp) variants, in resting and hyperpolarized states. The 3D-models were graphically inspected with PyMOL (Molecular Graphics System, version 1.8, Schrödinger, LLC) to analyse the impact of each variant on HCN2 structure. The solvent accessible surface area (ASA) and the relative solvent accessibility (RSA) were calculated with PyMOL with a solvent radius of 1.4 Å. HCN2 structures were also analysed using two predictive and analytical tools developed by Biosig Lab available at https://biosig.lab.uq.edu.au/. The tools called DDMut (https://biosig.lab.uq.edu.au/ddmut/) and DynaMut (https://biosig.lab.uq.edu.au/dynamut/) were used to compute the Gibbs free energy (ΔΔG) Δ vibrational entropy energy between wt-HCN2 and variant (ΔΔSVib ENCoM), giving information on structure flexibility.^39,40^

##### Graphs and statistical analysis

All graphs and statistical analyses were performed using GraphPad Prism□7.02 software (La Jolla, CA, USA). Data are mean ± standard error of the mean (SEM). The normality of sample distribution was assessed with the Shapiro-wilk test and, as appropriate, differences between groups were analysed with parametric or nonparametric tests, as indicated in the legends of the figures. The significance of the tests was considered for the following conditions: *P*<0.05 (*), *P*<0.01 (**), *P<*0.001 (***) and *P<*0.0001 (****).

### Data availability

Supporting data for the results of this study are available from the corresponding authors [CH, CL], upon reasonable request.

## Results

### Clinical characteristics of *HCN2*-related neurodevelopmental disorders

The clinical data of the twenty-one individuals (11 females, 10 males) reported here, whose ages ranged from 2 to 61 years (median age 8 years and 2 month), are summarized in Table 2. Developmental delay (DD/ID) was present in 17/21 individuals, and was of variable severity: mild (3/17, 18%), moderate (3/17, 18%), severe to profound (10/17, 59%), unspecified (1/17, 6%). Developmental regression was reported in four individuals, and was temporally related to epilepsy in one.

Twelve individuals (57%) had seizures, two having febrile seizures (FS) only and ten (48%) having epilepsy. The median age of epilepsy onset was 10 months (range 1 month - 6 years). A range of seizure types and EEG patterns were observed. One individual had infantile epileptic spasms syndrome, one had epileptic encephalopathy with spike-wave activation in sleep (EE-SWAS), and the remaining patients had unclassified epilepsies. No consistent treatment response was identified. Seizures were ongoing at last review in 3/10 individuals.

Tone abnormalities (hypo- or hypertonia) were present in sixteen individuals (76%) (Table 2). Ten individuals had movement disorders, including dystonia (n = 6), stereotypies (n = 5), static cerebellar signs (n = 1) and tremor (n = 2). Nystagmus was present in eight individuals (38%). Thirteen individuals (I1,I9-I11, I20-21) exhibited normal brain MRI. Abnormal brain MRI were observed for 9 individuals (I2, I5-8, I13-15, and I19). Brain MRI showed periventricular white matter T2 hyperintensities in five individuals, including the three carrying the p.(Glu478del) variant (Table 3, Supplementary Fig. 2).

### Identification of monoallelic and biallelic variants in *HCN2*

We identified thirteen different variants in the coding sequence of *HCN2*, twelve were novel, including eleven missense variants, one recurrent in-frame deletion (I13-15), one frameshift variant. The distribution of the HCN2 variants along the protein sequence is shown in Fig. 1A. Among these variants, seven were monoallelic including five *de novo*, one not maternally inherited and one inherited over three generations (Fig. 1B). The remaining six variants were biallelic including four homozygous and two compound heterozygous. Noteworthy, the compound heterozygous variants resulted from the unusual combination of one *de novo* variant (p.(Ser409Leu)) and one variant in trans (p.(His205Gln)), inherited from the asymptomatic mother. The p.(Glu478del) variant was found recurrently in three unrelated individuals, and two variants, namely p.(Arg324Cys) and p.(Arg324His), involved the same amino acid (AA). The p.(Gly460Asp) variant was previously described as a DEE causing variant.^28^ All thirteen variants have *in silico* prediction scores in favour of a deleterious effect. They all affect an AA that is highly conserved across all HCN subtypes in humans and other species (Fig. 1B), and are absent or extremely rare in the gnomAD database (Supplementary Table 1). Moreover, they all are clustered in the S4-S5 segments, the C-linker and the CNBD with no obvious correlation with the mode of inheritance.

### Electrophysiological studies of the monoallelic variants, p.(Arg324His), p.(Ala363Val) and p.(Met374Leu)

Next, we characterized three monoallelic variants by TEVC: p.(Arg324His), located in S4, and p.(Ala363Val), and p.(Met374Leu), located in S5 (Fig. 1A). With control non-injected oocytes, the observed currents were rectangular, strictly proportional to the stimulation intensity and did not exceed 0.72±0.10 nA/nF (maximal current density at Vm = −130 mV, n = 10), indicating the absence of voltage-gated endogenous currents activated by the voltage step stimulation protocol (Fig. 2A, B). The current families generated by the wt/p.(Arg324His) variant appeared similar in shape to that of wt-HCN2, but with much larger amplitudes (Fig. 2A). In contrast, the current families of wt/p.(Ala363Val), and wt/p/(Met374Leu) mixtures exhibited smaller amplitudes and different patterns (Fig. 2A). HCN2 currents exhibited three parts: an instantaneous step, a slowly activating phase that reaches a steady-state (Fig. 2A), as already described.^41^

**Figure 2.**
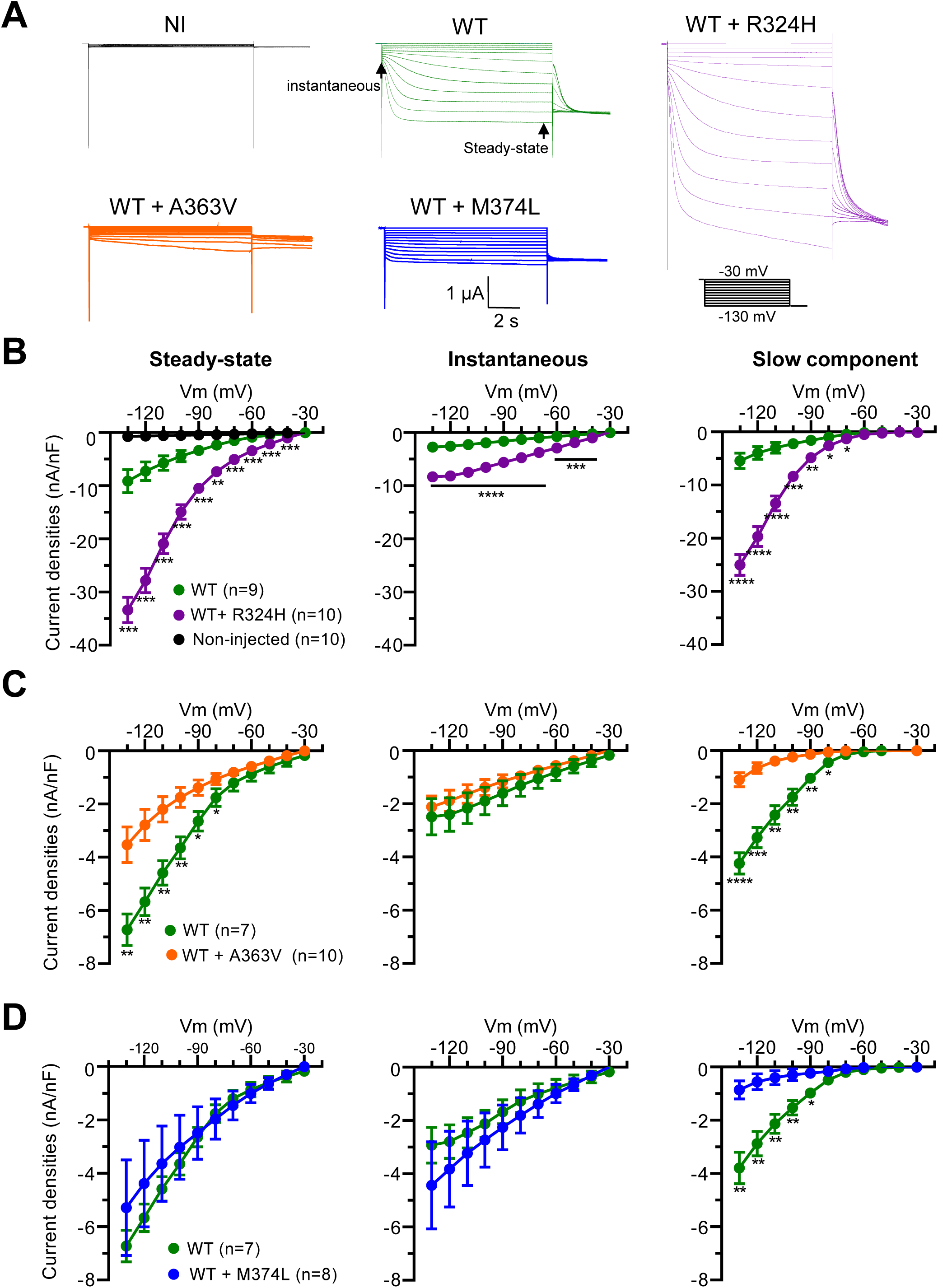
Electrophysiological characterization of wt-HCN2 and the monoallelic variants, p.(Arg324His), p.(Ala363Val) and p.(Met374Leu). **(A)** Examples of superimposed current traces, representing the current families, developed in response to hyperpolarized stimulations. From left to right, example of currents elicited by uninjected oocytes (NI), wt-HCN2 channel (WT), wt/p.(Arg324His) (WT + R324H), wt/p.(Ala363Val) (WT + A363V) and, wt/p.(Met374Leu) (WT + M374L) channels, respectively. **(B to D)** From left to right, the I/V curves were built using current densities measured at the steady-state and instantaneous components. Current densities for the slow component were then deduced. For each variant, significance of difference with wt-HCN2 was analysed with a two-way ANOVA test (corrected with Geisser-Greenhouse method), followed by a comparison test (two-stage step-up method of Benjamini, Kriger and Yekutieli). Data are shown as mean ± SEM. For clarity, no information indicates no significance. *, *P<*0.05; **, *P<*0.01; ***, *P<*0,001; ****, *P<*0.0001.

To characterize the impact p.(Arg324His), p.(Ala363Val) and p.(Met374Leu) variants, we compared the current densities of these three components with those elicited by wt-HCN2. Concerning wt/p.(Arg324His), the current densities of steady-state, instantaneous and slow components were significantly higher (3 to 5-fold) than those of wt-HCN2 from −130 to −70 mV (Fig. 2B). Concerning wt/p.(Ala363Val) and wt/p.(Met374Leu) combinations, we observed profound modifications of current shape in comparison with wt-HCN2 (Fig. 2A). The instantaneous components elicited by these two variants were similar to those of wt-HCN2 (Fig. 2C and D, middle graphs). The wt/p.(Ala363Val) variant elicited significantly lower current densities for both steady-state and slow components than wt-HCN2 from −130 to −80 mV (Fig. 2C, left and right graphs). For the p.(Met374Leu) variant, we observed lower current densities of the slow component only (Fig. 2D, right graph). Then, wt/p.(Ala363Val) and wt/p.(Met374Leu) variants induced a five and three-fold decrease of the slow component of HCN2 currents, respectively. The Western blots did not show any obvious significant protein reduction for either variant (Supplementary Fig. 3).

The slow component of wt-HCN2 and wt/p.(Arg324His) current traces could be best-fitted with a two exponential equation, whereas those of both wt/p.(Ala363Val) and wt/p.(Met374Leu) variants were best fitted with a mono-exponential equation, which was therefore used to analyse all traces (Fig. 3A). The current traces of p.(Arg324His) variant exhibited slightly faster activation kinetics only from −130 to −110 mV (*P<*0.05, Fig. 3B, left graph). For both wt/p.(Ala363Val) and wt/p.(Met374Leu) variants, we observed a significant increase in the rate of activation from −130 mV to −80 mV than of wt-HCN2, which was all the more important as Vm was positive (Fig. 3B, middle and right graphs).

**Figure 3.**
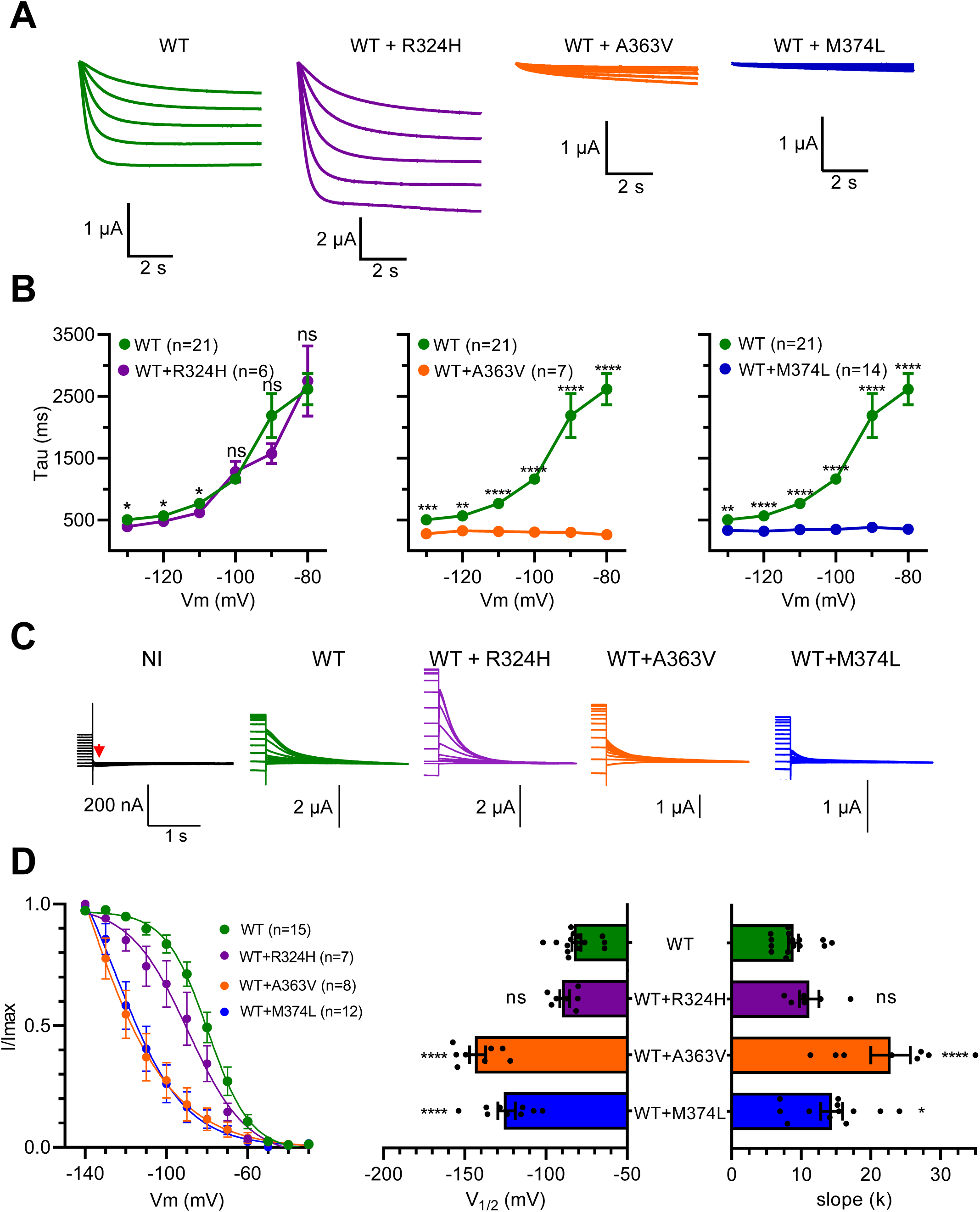
Activation kinetics and tail current analysis for wt-HCN2 and the monoallelic variants, p.(Arg324His), p.(Ala363Val) and p.(Met374Leu). **(A)** Examples of traces showing the slow component of currents elicited by wt-HCN2 channel (WT), wt/p.(Arg324His) (WT + R324H), wt/p.(Ala363Val) (WT + A363V) and, wt/p.(Met374Leu) (WT + M374L) channels, respectively. **(B)** Current kinetics analysis. Current traces were well-fitted with a mono-exponential equation. Tau (ms) was plotted versus Vm (mV). For each variant, significance of difference with wt-HCN2 was analysed with a two-way ANOVA test (corrected with Geisser-Greenhouse method), followed by a comparison test (two-stage step-up method of Benjamini, Kriger and Yekutieli). **(C)** Representative tail current traces for wt-HCN2 channel (WT), wt/p.(Arg324His) (WT + R324H), wt/p.(Ala363Val) (WT + A363V) and, wt/p.(Met374Leu) (WT + M374L) channels, respectively. **(D)** Left panel shows normalized current amplitudes (I/Imax) versus Vm were well-fitted with the Boltzmann equation, allowing the determination of the half-activation potentials (V1/2) and the slope factor k. Right panel shows the scatter plots show the comparison of the V1/2 (left) and slope factor (right) of the wt-HCN2, wt/p.(Ala363Val), wt/p.(Met374Leu) and wt/p.(Arg324His) mutant channels. A one-way ANOVA test (F = 53,15, *P<*0.0001), followed by a Dunnett test, was performed and showed a significant difference between mutants and wt-HCN2, except between p.(Arg324His) and wt-HCN2 (p-value = 0.48). Data are shown as mean ± SEM. ns, not significant; *, *P<*0.05; **, *P<*0.01; ***, *P<*0,001; ****, *P<*0.0001.

Next, we analysed the instantaneous tail currents that correspond to activation phase, using the Boltzmann equation (Fig. 3C). The data were well fitted, allowing the determination of the half potential of activation (V_1/2_) and the slope coefficients (k). With wt/p.(Arg324His) channel, the V_1/2_ and k-values were not significantly different from those of wt-HCN2 channel (Fig. 3D). In contrast, with both wt/p.(Ala363Val) and wt/p.(Met374Leu) channels, V_1/2_ of activation were shifted to the left by 60.8 and 43.0 mV, respectively (*P<*0.0001, Fig. 4D). This hyperpolarized shift was significantly larger for p.(Ala363Val) variant than p.(Met374Leu) variant (*P<*0.0001, Fig. 3D, middle graph). For these two variants, the slopes were significantly shallower than that of wt-HCN2 (*P<*0.0001, Fig. 3D, right graph).

**Figure 4.**
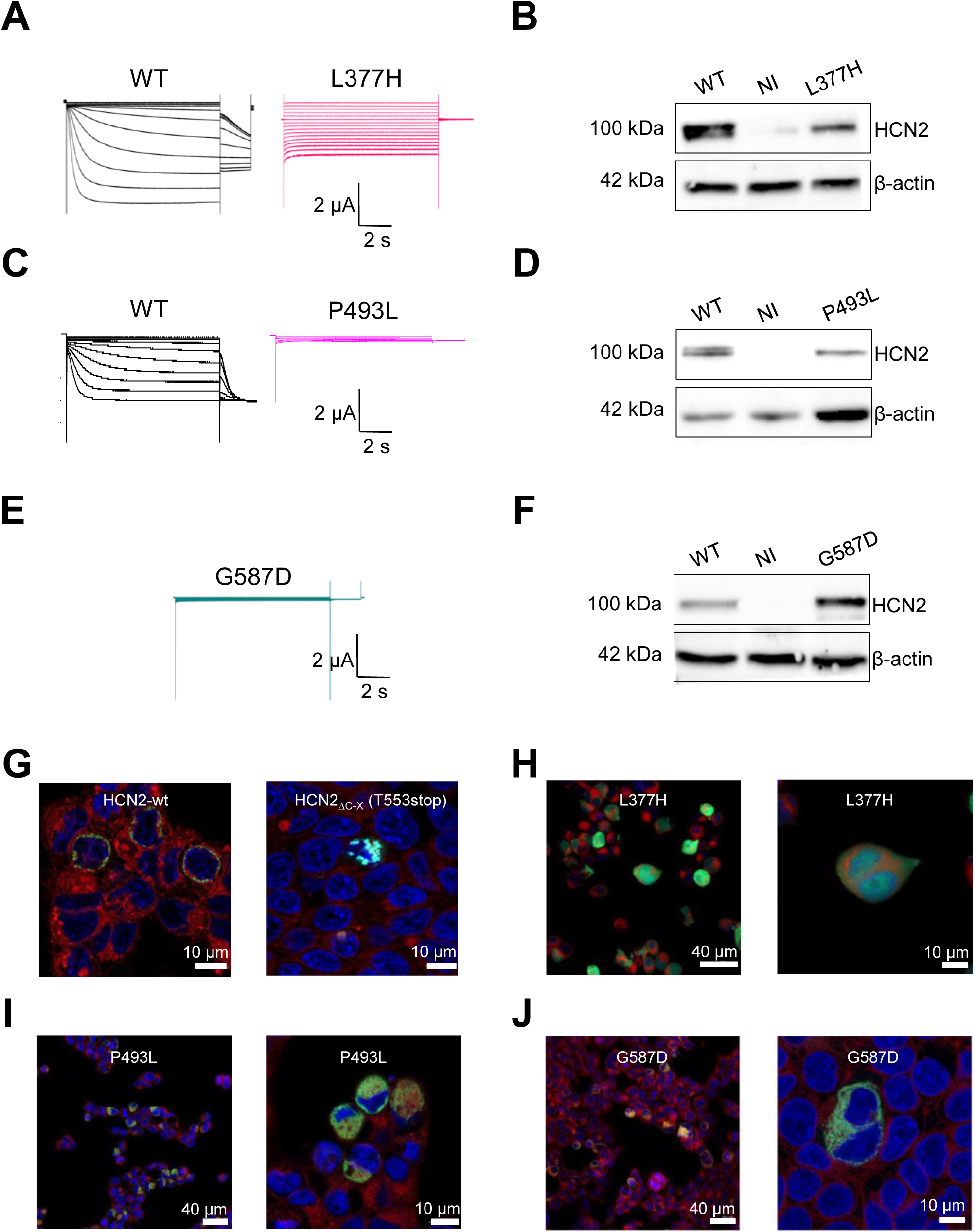
Characterization of p.(Leu377His), p.(Pro493Leu) and p.(Gly587Asp) HCN2 variants. **(A, C and E)** Representative TEVC recordings from oocytes expressing HCN2 wild-type and two homozygous variants of HCN2, p.(Leu377His), p.(Pro493Leu) and p.(Gly587Asp). The data show current traces for voltage steps from −130 to −30 mV from a holding potential of −30 mV for 10 s. **(B, D and F)** Western blot of *Xenopus laevis* oocyte proteins. The panels show immunostaining with anti-HCN2 and anti-β-actin antibodies. WT: oocytes injected with wt-HCN2 mRNA, NI: non-injected oocytes; L377H, P493L and G587D: oocytes injected with p.(Leu377His), p.(Pro493Leu) and p.(Gly587Asp) variants mRNAs, respectively. **(G-J)** Study of membrane trafficking of p.(Leu377His), p.(Pro493Leu) and p.(Gly587Asp) variants in HEK293 cells. The expression of EGFP-tagged p.(Leu377His) or p.(Pro493Leu) or p.(Gly587Asp) HCN2 variants was evaluated by confocal microscopy. The cells were stained with the CellMask orange membrane stain and DAPi nuclear stain. EGFP-tagged wt-HCN2 **(G,** left panel) shows a strong EGFP signal at the plasma membrane, while the trafficking-defected HCN2Δ_C-X_ (Thr553Ter) shows cytosolic staining **(G,** right panel), as seen for p.(Leu377His), p.(Pro493Leu) and p.(Gly587Asp) variants **(H-J)**. Examples of images obtained from magnification of 63x are shown **(G and H-J, left panels)**. Examples of overview with magnification from 20x are also shown **(H-J, left panels)**.

Taken together, our electrophysiological data show that the p.(Arg324His) variant results in GOF, whereas the p.(Ala363Val) and p.(Met374Leu) variants induce a partial LOF, when they are expressed in *Xenopus* oocytes.

### Electrophysiological, Western blot and immunocytochemical analyses of the biallelic variants, p.(Leu377His), p(Pro493Leu) and (Gly587Asp)

The biallelic variants p.(Leu377His), p.(Pro493Leu) and p(Gly587Asp), were then individually expressed in *Xenopus* oocytes to investigate their functional impact on HCN2. The data from expression experiments of p.(Leu377His), p.(Pro493Leu) and p.(Gly587Asp) are shown in Fig. 4. Neither p.(Leu377His) (n = 8), p.(Pro493Leu) (n = 6) nor p.(Gly587Asp) (n = 4) produced any measurable currents (Fig. 4A, C and E). Western blot showed a strong immunoreactivity for two bands with apparent molecular weight close to 100 kDa with wt-HCN2 (Fig. 4B, D and F). These two bands likely reflect a glysosylated and a non-glycosylated form of wt-HCN2.^38^ However, p.(Leu377His) and p.(Pro493Leu) showed only one band corresponding to the non-glycosylated form, while p(Gly587Asp) variant behaved as wt-HCN2. We hypothesized that the LOF of these three *HCN2* variants could be the consequence of the alteration of membrane trafficking. We addressed this question by expressing p.(Leu377His), p.(Pro493Leu) and p(Gly587Asp) variants tagged with EGFP in HEK293 cells and imaging their trafficking to the plasma membrane by confocal microscopy (Fig. 4G-J). While EGFP-tagged wt-HCN2 showed staining of the plasma membrane (Fig. 4G), no obvious membrane expression was observed for p.(Leu377His), p.(Pro493Leu) and p(Gly587Asp) variants (Fig. 4H-J). Indeed, a strong EGFP-staining of cytosol was observed, indicating that all biallelic variants remain in the cytosol. In conclusion, these data evidence that the biallelic p.(Leu377His), p.(Pro493Leu) and p.(Gly587Asp) variants induce a complete LOF, due to the impairment of membrane trafficking.

### Structural Impacts of the identified pathogenic variants onto a 3D Homology model of HCN2

Next, we built 3D homology models of the HCN2 channel to further investigate the structural impact of all variants reported in this work (Fig. 5). We thus examined each mutation in both depolarized and hyperpolarized states. We first examined the effects of each missense mutation on HCN2 structure stability, by analysing the ΔΔG values (Table 4). The idea was also to check whether the prediction of the pathogenicity of these variants fit with the clinical and electrophysiological data. The DDMut server indicated that p.(His205Gln)/p.(Ser409Leu), p.(Leu377His), p.(Pro493Leu), and p(Gly587Asp) variants induced destabilizing effects on HCN2 structures at both depolarized and hyperpolarized states. Interestingly, high destabilizing effects were determined for p.(His205Gln)/p.(Ser409Leu), p.(Pro493Leu), and p.(Gly587Asp) variants and p.(Gly460Asp) variant has a very weak stabilizing effect. In contrast, p.(Arg324His), p.(Arg324Cys), p.(Ala363Val), p.(Asn369Ser) and p.(Met374Leu) variants show positive ΔΔG values, indicating stabilizing effects on HCN2 structure (Table 4). Thus, we carefully examined the 3D structure of each HCN2 variant 3D model structure to identify possible structural damage induced by these variants.

**Figure 5.**
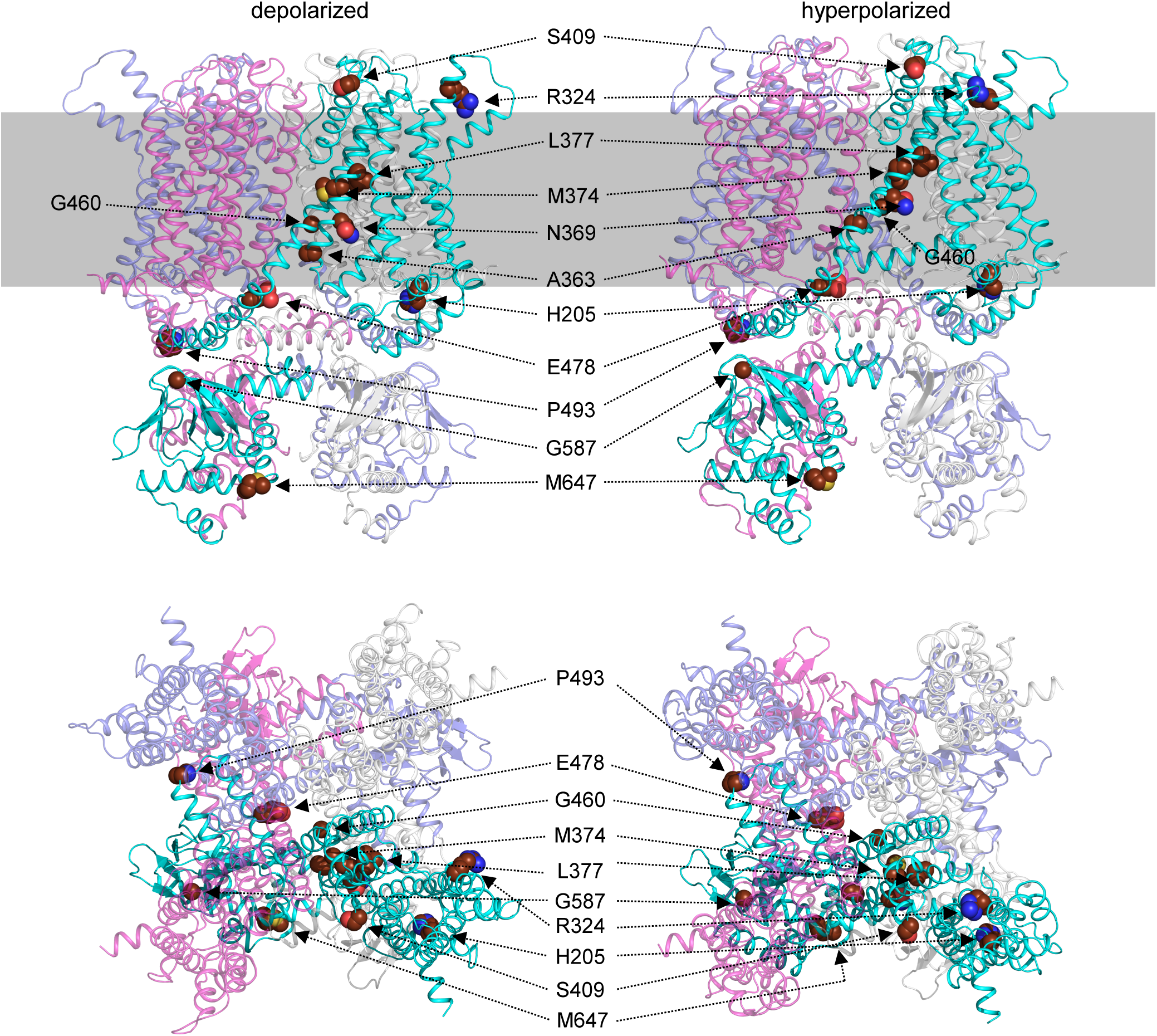
Mutation mapping onto 3-dimensional homology models of HCN2 in depolarised and hyperpolarised states. Models of HCN2 in depolarized (left panels) and hyperpolarized states (right panels) are shown (upper panels: profile views. lower panels: top views). Proteins are drawn in ribbon representation with a different color code for each subunit. For clarity, the locations of mutated amino acids are represented as spheres in only one subunit (in cyan), while the other subunits are shown in semi-transparent representations. The approximate position of HCN2 within the membrane is visualized with the gray rectangle.

The p.(Arg324His) and p.(Arg324Cys) variants concern the second positively charged residue of S4. In the depolarized state (or close conformation), Arg324 side chain is oriented towards the extracellular compartment and thus freely exposes its positive charge. Interestingly, during HCN2 opening, S4 rotates counterclockwise, burying the Arg324 side chain in a hydrophobic pocket which is composed by Ile233, Leu323, Tyr317, Leu323, Phe328, Met391 and Leu391 (Fig. 6A). The ASA values of Arg324 dropped from 139.27 Å^2^ (RSA = 64%) to 24.07 Å^2^ (RSA = 14.12%) between depolarized state and hyperpolarized state (Table 4). In p.(Arg324His) variant, the shorter side chain of His, at position 324, fitted better in this hydrophobic pocket in the hyperpolarized state (ASA = 14.12 Å^2^, RSA = 7%) and could be stabilized by π-interaction with Tyr317 and Phe328 (Fig. 7B, lower left panel). In p.(Arg324Cys), Cys, which is more hydrophobic than Arg and His, can also be more stabilized in the hydrophobic pocket than Arg (ASA = 9.44 Å^2^, RSA = 7%) (Fig. 6B, lower right panel).

**Figure 6.**
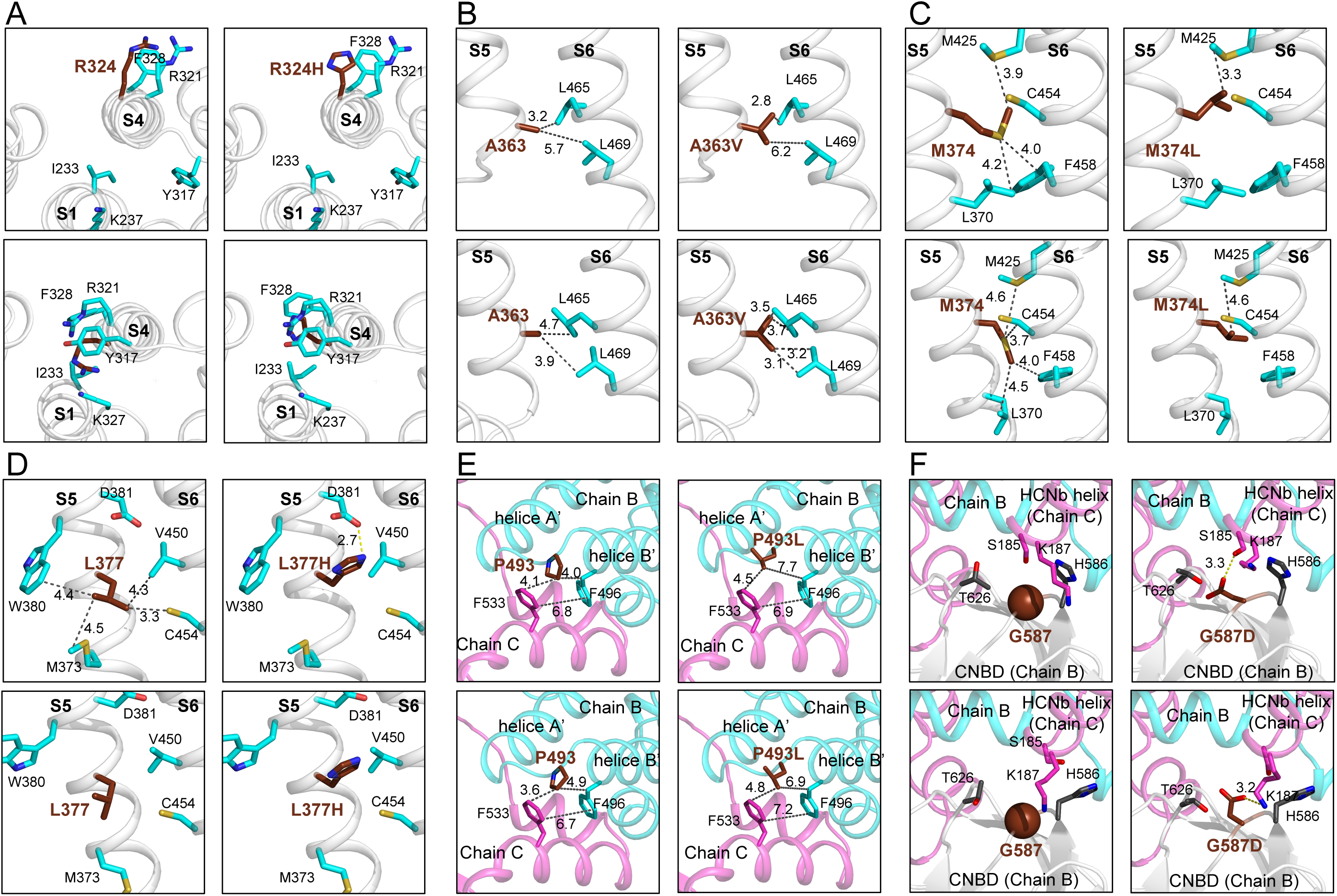
Structural analysis of p.(Arg324His), p.(Ala363Val), p.(M374Leu), p.(Leu377His), p.(Pro493Leu) and p.(Gly587Asp) HCN2 pathogenic variants. For clarity, zoom in of profile views were chosen for each mutation, except for p.(Arg324His) variant. **(A)** Top view of S1-S4 helices of wt-HCN2 and p.(Arg324His) HCN2 variant in the depolarized and hyperpolarized states. The side chain of residue at 324 is towards the solvent in the depolarized state of HCN2 (left panel), but after opening it remains trapped in a hydrophobic pocket formed by I233, L323, Y317, F328, M390 and L391. **(B, C and D)** Profile views of S5-S6 interfaces showing the residues A363 **(B)**, M374 **(C)** and L377 **(D)**. **(E and F)** The two last mutations are located in the intracellular region of HCN2 structure at the interfaces between two subunits (Chains B and C). The p.(Pro493Leu) variant **(E)** might destabilize the bend between helices A’ and B’ and also the N-terminal of helice B’. The p.(Gly587Asp) variant could change the curvature of the loop between two β-sheets within the CNBD **(F)**. The proteins are drawn in ribbon representation. and their closest-neighbour contacts with residues of S6. The distances are indicated in Å.

The next variant, p.(Ala363Val) involves important changes in the N-terminus of α-helix S5, which impacted hydrophobic interaction with two leucines (Leu465 and Leu 469) in α-helix S6, which are in the nearest-neighbour of Ala363 (Fig. 6C). During channel opening, the side chain of Ala363 is closer to these residues. At this position, Val, which is bulkier and more hydrophobic, could form stronger hydrophobic interaction with Leu465 and Leu469 (Fig. 6C, right panels). These two leucines are even closer to Ala in the hyperpolarized state (Fig. 6C, lower right panel). This could affect the stability of the hyperpolarized state of HCN2 and might accelerate its activation kinetics, in agreement with the positive value of ΔΔG identified with DDMut (Supplementary Table 5). In addition, the p.(Aal363Val) variant induced a decrease of molecule flexibility (with ΔΔSVib ENCoM values of −0.626 and −0.287 kcal.mol^-1^.K^-1^, respectively) at both depolarized and hyperpolarized states.

In the depolarized state of HCN2, the side chain of Met374 is in sandwich between Phe458 and Met425 of α-helix S6, while in the hyperpolarized, it goes between Phe458 and the backbone of Cys454 (Fig. 6D, left panels). In p.(Met374Leu) structure, a Leu at position 374 exhibits a different orientation with stronger interaction with Met425. This might strongly impact these interactions and involves important changes in the interface between α-helix S5 and S6, and consecutively HCN2 opening (Fig. 6D, right panels). This is supported by the increase of ΔΔG values, and reorganization of the S4-S5 segment during channel opening already reported.^42^ Moreover, the p.(Met374Leu) variant induced an increase of flexibility of the molecule at the depolarized state (ΔΔSVib ENCoM = −0.309 kcal.mol^-1^.K^-1^), while this mutation led to increased rigidity at the hyperpolarized state (ΔΔSVib ENCoM = 0.063 kcal.mol^-1^.K^-1^).

In the depolarized state of wt-HCN2, Leu377 makes strong hydrophobic interactions with Val450 and Cys454, at the interface between S5 and S6 (Fig. 6D, upper left panel). Apparently, these interactions are absent in the hyperpolarized state, suggesting that the opening breaks them (Fig. 6D, lower left panel). In the depolarized state of p.(Leu377His) variant, the side chain of His exhibits a different orientation than Leu and could strongly interact with Asp381 through a salt bridge (Fig. 6D, upper right panel). In addition, with His at position 377, all hydrophobic interactions at the S5 and S6 interface are lost. Thus, the structural modifications induced by a His at this position might explain the LOF of HCN2 and account for the large leak currents associated with this construct (Fig. 4A).

In the C-linker of HCN2, the p.(Pro493Leu) and p.(Glu587Asp) variants modify residues which are buried and important for the structure and dynamics of HCN2. Since Pro493 stabilizes the kink between the A’ and B’ anti-parallel helices by interacting with Phe533 of a neighbour subunit, the Pro493Leu mutation should thus strongly destabilize the channel structure (Fig. 6E). Gly587 is located at a central turn of the CNBD β core, in the vicinity of Lys187 of a neighbour subunit (Fig. 6F). Its substitution by an aspartic acid should lead to a salt bridge between the CNBD and the channel N-terminus. In addition, the Phi/Psi angles are outlier with G587D mutation.

We also performed a structural analysis of p.(His205Gln)/p.(Ser409Leu), p.(Asn369Ser) and p.(Gly460Asp) variants and highlighted their structural differences with wt-HCN2. The trans variant p.(His205Gln))/p.(Ser409Leu) variant showed profound structural changes (neutralisation of a buried charge, disruption of salt bridge between His205 and Glu192 in helix HCNc and loss of aromatic interaction between H205 and F183 of helix HCNc and Y207 (Supplementary Fig. 4A). Thus, we assumed that the trans variant p.(His205Gln)/ p.(Asn369Ser) could have a deleterious impact on HCN2 function. The p.(Asn369Ser) variant leads to the loss of a hydrogen bound with Glu351 in S4 (Supplementary Fig. 5A). Finally, the p.(Gly460Asp) variant impacts the interface between S6 from two adjacent subunits, leading to clash with Ala462, Thr463 and Ile466 (Supplementary Fig. 6).

### Phenotypic differences between individuals with GOF and LOF variants

In individuals carrying a GOF variant (n = 18, including the 3 individuals from Family 2 and 15 individuals in the literature), seizures were present in 16/18 (89%) (FS only in 8/16, epilepsy in 8/16) with median age of onset of 5 years, but intellectual disability was present in only 1/18 individuals (5.5%) (Table 2). In individuals carrying LOF variants (n = 12, being individuals 6, 7, 9, 10, 11, 12, 16, 17, 18, 19 from this cohort and two individuals from literature), epilepsy was present in 8/12 (67%) with median age of onset of 7.5 months. ID was present in 11/12 (92%) individuals (not stated in one individual), being of moderate severity in two individuals with monoallelic LOF and severe-profound in three individuals with biallelic LOF. Of the 12 individuals whose variants have not been studied functionally, the six with biallelic variants all had severe DD/ID (Table 2).

## Discussion

In this study, by describing twenty-one additional individuals with deleterious *HCN2* variants, we expand the phenotype of *HCN2*-related disorders, from mild epilepsies with normal cognition, to phenotypes, including ID with or without epilepsy, and individuals with DEE. We reported twelve novel *HCN2* variants and highlight by combining electrophysiological and trafficking data with 3D-modelling that these variants can cause either GOF (monoallelic variants) or partial LOF (monoallelic variants) or complete LOF (biallelic variants). Taken together, we provide preliminary evidence of phenotype-genotype correlation: specifically, that LOF variants may be associated with more severe ID and, where epilepsy is present, earlier age of seizure onset.

We provide comprehensive electrophysiological and structural analysis to demonstrate that some variants lead to loss of HCN2 channel function. Our electrophysiological data clearly demonstrated that p.(Ala363Val) and p.(Met374Leu) variants strongly impair HCN2 operation, but still exhibit the main features of an hyperpolarized-activated ion channel. Indeed, both the p.(Ala363Val) and p.(Met374Leu) variants cause a strong decrease of current densities and a visible alteration of current kinetics. The negative shift of activation properties of both variants also indicates an impairment of the voltage-dependency. Thus, we attribute these alterations to a partial LOF of HCN2. This assumption is reinforced by the examination of our 3D-model, suggesting modification in the interactions of Ala363 and Met374 with Leu465 and Phe468 in S5, respectively. Altogether, we hypothesize that p.(Ala363Val) and p.(Met374Leu) variants alter the interactions between the S4, containing the voltage sensor and S5. The detrimental effects of mutations in the S4-S5 regions of tetrameric K_V_ and related HCN channels, is increasingly well documented with reports of cation leakage and gating blocks.^23,43,44^ Interestingly, the p.(Met305Leu) HCN1 variant, which is an homologous variant to the p.(Met374Leu) variant has been associated with DEE.^44^ While the p.(Met 374Leu) HCN2 variant shows similar current kinetics to the p.(Met305Leu) HCN1 variant, the instantaneous tail currents elicited by both variants shift in opposite direction.^22^ This could be explained by biophysical specificities of HCN1 versus HCN2. In conclusion, p.(Ala363Val) and p.(Met374Leu) variants likely cause a partial LOF of HCN2, which could impair neuronal excitability.

In contrast, p.(Leu377His), p.(Pro493Leu) and p.(Gly587Asp) variants produce HCN2 proteins which are electrophysiologically silent in *Xenopus* oocytes. The p.(Leu377His) would impair the opening of HCN2 through the loss of the interaction with Gly450 in S6. The p.(Pro493Leu) variant could strongly destabilize the channel structure through the alteration of the interaction of the kink between the A’ and B’ anti-parallel helices of a neighbour subunit. The p.(Gly587Asp) variant located in the CNBD, would not influence cyclic-nucleotide binding, but the aspartate could form a salt bridge with Lys187 in the N-terminal HCN domain, disrupting channel gating.^45^ In addition, these three variants could not be detected at the plasma membrane, indicating that p.(Leu377His), p.(Pro493Leu) and p.(Gly587Asp) variants impair HCN2 membrane trafficking. To date, only two variants that alter membrane trafficking in HCN channels family have been reported, one in in HCN4 p.(Pro257Ser) and one in HCN2 p.(Gly460Asp), associated with early-onset atrial fibrillation and DEE, respectively. ^28,46^ We assumed that the trafficking-impairment p.(Leu377His) variant might be the consequence of the misfolding of HCN2, while p.(Pro493Leu) and p.(Gly587Asp) variants could affect the interaction with trafficking regulating proteins, such as TRIP8b.^35^ Altogether, p.(Leu377His), p.(Pro493Leu) and p.(Gly587Asp) variants are leading to complete LOF by impairing membrane trafficking.

Our study brings novel evidence to clearly establish LOF as a mechanism of HCN2-related disease, building on two previous case reports. As recently suggested by DiFrancesco, who reported that the p.(Gly460Asp) results in LOF, we believe that the pathogenicity of monoallelic LOF HCN2 variants is due to a dominant negative effect rather than haploinsufficiency, as evidenced by three observations.^28^ Firstly, in our cohort (seven cases from four families) and the literature (one case), the heterozygous parents of a child with a homozygous biallelic variant were all unaffected. Secondly, disease-causing monoallelic truncating HCN2 variants have not been reported to date; similarly, truncating pathogenic variants have not been reported in *HCN1* or *HCN4*. Thirdly, data from the gnomAD database includes twelve truncating *HCN2* variants in healthy individuals, and a pLI score of 0.17 indicating a low probability that *HCN2* could be intolerant to loss of function.

We also provide data to advance understanding of GOF *HCN2* variants. The p.(Arg324His) variant led to an increase of HCN2 ion channel function as the result of an increase of conductance and faster activation kinetics compared to wt-HCN2, while voltage-dependent activation properties are unchanged. In addition, the 3D-model did not reveal any modification at the molecular level in the depolarized state. However, in the hyperpolarized state, the side chain of the residue at position 324 falls into a hydrophobic pocket, which constitutes a more stabilizing environment for histidine and cysteine than for the arginine. The p.(Arg324His) variant could generate a lower electrostatic repulsion of cations at the extracellular side of the channel, which favors cations to flow through HCN2. Further molecular dynamic analysis will be required to better understand the impact of these mutations. Thus, the GOF caused by p.(Arg324His) variant results from an important increase of HCN2 conductance as reported for the p.(Pro719-Pro721del) variant, but also from faster activation kinetics, not seen for the p.(Pro719-Pro721del) variant (Table 1).^24^ Two other monoallelic variants of *HCN2* (p.(Val246Met) and p.(Ser635Trp)) with GOF have been already described, while these variants exhibit unchanged conductance, they give faster currents and activation at less negative potential than wt-HCN2.^27^ Overall, these findings highlight that, similar to other ion channelopathies, GOF can arise from disruption of a number of different aspects of HCN2 channel function.

With this cohort, we confirm that pathogenic *HCN2* variants can be associated with more severe phenotypes than previously reported.^24–27^ To date, only six variants – p.(Pro719-Pro721del), p.(Val246Met), p.(Ser632Trp), p.(Glu515Lys), p.(Ser126Leu) and p.(Gly460Asp) – of *HCN2* have been reported in 18 individuals with relatively mild forms of epilepsy including GGE and GEFS+ and most of them had no ID.^24–28^ In contrast, seventeen out of the twenty-one individuals reported here have mild to profound ID and only ten have a history of seizures. The epilepsy phenotypes in the individuals in our cohort are variable, and most do not have features consistent with a named epilepsy syndrome. Abnormal tone, movement disorders, particularly dystonia and stereotypies, and nystagmus are common symptoms, being present in 76%, 48% and 38% of individuals respectively; all these neurological findings broaden the phenotypic spectrum associated with *HCN2.* Five individuals have abnormal brain imaging, showing similar patterns of bilateral periventricular or subcortical white matter hyperintensities; this pattern is not reported in the remaining individuals. We did not identify other common symptoms in the group, although acknowledge that some may have been under-recognized or under-reported.

Similar to *HCN1*, our data and the literature allowed us to highlight limited genotype-phenotype correlation for HCN2-related conditions (Table 1). Seizures were a common but not universal feature in those with either GOF or LOF variants, being present in 89% (16/18) and 67% (8/12) respectively. However, only 44% (8/18) individuals with GOF variants had epilepsy, the remaining individuals having FS only. Additionally, the age of epilepsy onset was earlier in those with LOF variants (median 7.5 months) compared with GOF variants (median 5 years). DD/ID was present more frequently and was more severe in those with LOF variants (present in 11/12 individuals (92%), range moderate-severe) than with GOF variants (present in 1/18 individuals (6%), range normal-mild). We note that features other than the *HCN2* variant likely contribute to disease severity: we saw intrafamilial variability in severity for family 2 with the monoallelic p.(Arg324His) variant and for families 7, 11 and 14 carrying the homozygous p.(Leu377His), p.(Pro493Leu) and p.(Met647HisfsTer31) variants, respectively. Additionally, we noted interindividual variability for the two individuals carrying the monoallelic p.(Ala363Val) variant in terms of ID severity and presence of epilepsy; and the three individuals carrying the inframe p.Glu478del variant, with variability in severity of visual impairment and movement disorder.

Taken together, our data indicate that the related HCN2 phenotype, ranging from febrile seizures or mild epilepsy without ID, to severe forms of epilepsy with ID and ID without epilepsy, is very similar to that caused by HCN1 variants.^23^ Thus, one characteristic shared by both isoforms *HCN1* and *HCN2* is the wide phenotypic spectrum caused by channel disruption. A relevant difference between *HCN1* and *HCN2* is the absence of reported biallelic pathogenic variants in *HCN1*, while biallelic inheritance is frequent in our HCN2 cohort. Interestingly, monoallelic and biallelic pathogenic variants have also been described for genes encoding other ion channels, such as *SCN1B* and *CACNA1D*, both of which have been implicated in severe forms of epilepsy.^47,48^

We acknowledge limitations of our work, including that we did not perform functional studies on all the identified *HCN2* variants, and therefore cannot confidently determine whether they result in GOF or LOF, although we would expect LOF for the biallelic variants. However, the structural analyses of the monoallelic variants - p.(Arg324Cys), p.(Asn369Ser) and p.(Glu478del) - the biallelic p.(Met647HisfsTer31) variant and the two heterozygous compound variants - p.(His205Gln) and p.(Ser409Leu) - suggest significant structural impacts and consequently HCN2 channel dysfunctions (Supp. Fig. 3). Furthermore, there remain some unanswered questions about the impact of each variant on channel function. As performed elsewhere, molecular dynamic simulation studies of the effect of these variants on HCN2 channel 3D-structure would provide additional information on the alteration of gating mechanisms.^23,49^ At a neuronal circuit and whole brain level, the implications of HCN2 channel dysfunction are yet to be elucidated. The use of stem cell and organoid models, and generation of transgenic mice, will advance understanding of disease mechanisms, and provide models upon which to develop targeted therapies, as done recently for *HCN1*.^50^

In conclusion, our findings highlight the association of mono (n = 7) and bi-allelic (n = 6) *HCN2* variants with ID with or without epilepsy. To date, the p.(Leu377His), p.(Pro493Leu) and p.(Gly587Asp) variants represent the first cases of bi-allelic *HCN2* variants causing a complete LOF. Finally, these data are strongly supported by electrophysiological and structural results disclosing that both GOF and LOF variants are responsible for *HCN2*-related neurodevelopmental disorders.

## Data Availability

All data produced in the present study are available upon reasonable request to the authors

## Acknowledgements

We are very grateful to the patients and their family for their participation in this study. We would also like to thank the Groupama Foundation “Vaincre les maladies rares” for its financial support.

## Supplementary material

Supplementary material is available.

## Appendix 1

This section is included if the article contains an appendix, for example, to list consortium collaborators who must be indexed online.

## Abbreviations

AA: amino-acid
CNBD: cyclic nucleotide-binding domain
DEE: developmental and epileptic encephalopathy
EE-SWAS: epileptic encephalopathy with spike-wave activation in sleep
FS: febrile seizures
GEFS: generalized epilepsy with febrile seizures
GGE: genetic generalized epilepsy
GOF: gain-of-function
HCN: hyperpolarization activated cyclic nucleotide gated channel
I_max_: maximal current
ID: intellectual disability
LOF: loss-of-function
SOS: standard oocyte saline
TEVC: two-electrode voltage-clamp method
wt-HCN2: wild-type HCN2

