## Supplemental Material for "Mono and biallelic variants in *HCN2* cause severe neurodevelopmental disorders"

**Two-Electrode Voltage Clamp Recording in *Xenopus* oocytes**

Adult female *Xenopus laevis* were purchased from CRB (Rennes, France) or TEFOR (Paris-Saclay, France) and bred in the animal facility in strict accordance with the recommendations of the Guide for the Care and Use of Laboratory Animals of the European Community (European Community council directive 2010/63/EU). Oocytes were harvested from mature female *Xenopus laevis* frogs under 0.15% tricaine anaesthesia. All animals recovered after 2-3 h. Every female is operated every three months, not less. A single female was used no more than 5 times. The experimenters followed the steps to avoid any animals' pain and suffering, and they understand the ethical principles and state that their work complies with its animal ethics checklist. No statistical test was realized to estimate the sample sizes. Only one measurement was performed with each oocyte. Electrophysiological recordings were replicated from 4 to 12 distinct oocyte collected from at least three different frogs. When Ih exceed 400 nA at -30 mV, or when oocytes were leaky, recordings were excluded. Randomization was achieved by random selection of frog or oocytes before injection.

TEVC recordings were conducted using a TEV-200 amplifier (Dagan Corporation, Minneapolis, USA). Digidata 1440A interface (Axon CNS Molecular Devices, San Jose, CA, USA) and pCLAMP™ 10.7 software (Axon CNS Molecular

Devices) were used for current recording. Non-injected oocytes were used as negative controls and wt-HCN2 injected oocytes as positive controls. Injected oocytes were continuously perfused at room temperature with a standard oocyte saline solution (SOS) (in mM): 100 NaCl, 1 KCl, 1 $CaCl_2$, 1.8 $MgCl_2$, and 10 HEPES (pH = 7.5 using NaOH). Oocytes were impaled with electrodes filled with 0.5 M KCl/1.5 M K-acetate. A solution of $CsCl_2$ (5 mM in SOS) was used to block currents elicited by wt-HCN2 and its variants. For p.(Gly587Asp) and p.(Leu377His) variants, TEVC recordings were carried out in the Reid laboratory, Florey Institute of Neuroscience, Parkville, Australia using methods as described elsewhere.[1]

Currents were analysed using Clampfit 10.7 (Molecular Devices). Mean current amplitudes were calculated from at least five different cells from at least two different *Xenopus* females. Membrane capacitances were measured for each oocyte to determine current densities (nA/nF) which were used to analyse the current density-voltage relationship (I/V curves). I/V curves were plotted with steady-state and instantaneous currents. Slow currents were determined by subtracting instantaneous currents from steady-state currents. Current activation rates were analysed with classical exponential equation ($I = A.e^{-t/tau} + B$), where t corresponds to time, tau, the kinetics constant and B to the current when t = 0. For tail current or voltage-dependent activation analyses, we used the Boltzmann equation to determine half-activation potentials ($V_{1/2}$) and slopes (k).

**Western blot analysis**

Western blotting was performed as previously described[2], with initial centrifugation steps added to remove yolk proteins. Samples of 15-20 oocytes, injected or un-injected, were collected, resuspended in 15 µl/lysis buffer {20 mM HEPES buffer (pH 7.4) with protease inhibitors (Complete, Boehringer Mannheim)}, and lysed by multiple passages through a fine syringe needle. Yolk proteins were removed from the lysates by a centrifugation at 1000 g for 10 min. The pellet was discarded, and supernatant protein levels analysed by the Bradford method (Bio-Rad CA, USA). Supernatants were stored at -80°C. Prior to electrophoresis the samples were thawed, and the buffer adjusted to 20 mM HEPES (pH = 7.4), 1% Triton X100, 0.1% sodium deoxycholate, 1% SDS, 140 mM sodium chloride, 2 M urea (final concentration). The homogenates were then incubated with rotation for 30 min at 4°C to solubilize membrane proteins before 1x reducing loading buffer was added and the samples incubated at 37°C for 1 h.[3] Thirty µg of protein (corresponding to the protein content of one oocyte) from control and experimental samples were separated by electrophoresis on 8% SDS-PAGE gels, at 70-100 V, running buffer (Tris/glycine pH 8.3, 1% SDS) until the migration distance of 75 and 100 kDa size markers {(Pre-stained Precision Plus Dual Colour Standards (Bio-Rad, CA, USA)} reached approximately 3 cm. Proteins were transferred to nitrocellulose (Tris-glycine 1% SDS, 20% methanol), and controlled by Ponceau S staining

(Sigma Aldrich,#. P3504). Blots were blocked in 0.5% skim milk TBSI, 1x TBS 0.5% IGEPAL CA-630, (Sigma Aldrich, #I8896) for 1 h at RT. before overnight incubation at 4°C, with blocking buffer containing 1:500 rabbit anti-HCN2 APC-030 (Alomone Labs Cat# APC-030, RRID: AB2313726). The filters were washed with TBSI thrice and incubated in blocking buffer with goat anti-rabbit Poly HRP 32260 (Thermo Fisher Scientific Cat# 32260, RRID:AB1965959) 1:15,000 for 1 h at RT. Beta- actin (Thermo Fisher Scientific Cat# PA1-183, RRID:AB_2539914) was used as a loading control. The protein signal was visualized with Clarity Western ECL Substrate (Bio-Rad, CA, USA) and the signal captured by a Bio-Rad ChemiDoc™ MP imaging system (Image Lab Software, RRID: SCR_014210).

**Transient expression in HEK293 cells and confocal fluorescent microscopy**

HEK293T cells were transfected with N-terminal EGFP tagged constructs encoding human wt-HCN2, and the biallelic variants, p.(Leu377His), p.(Pro493Leu) and p.(Gly587Asp). The EGFP tagged and truncated $HCN2_{\Delta C\text{-}X}$ (Thr553Ter) was used as negative control. The HCN2 constructs were tagged at their amino termini to avoid disruption of membrane trafficking.[4] All reagents used for cell culture and transfection were purchased from Thermo Fisher Scientific (Scoresby, Victoria, Australia) unless otherwise mentioned. Transfections were carried out using Invitrogen Lipofectamine 3000 in 24 well plates containing glass coverslips (13 mm diam.) coated with poly D-lysine hydrobromide (Merck KGaA, Darmstadt, Germany), according to the manufacturer's instructions. In brief, 24 hours after splitting 1x $10^5$ cells/well, viability 99% were seeded in the medium, DMEM, high glucose (4.5 g/L), sodium pyruvate, no glutamine, supplemented with 10% GlutaMax Supplement, 10% fetal bovine serum, penicillin (100 U/ml) and streptomycin (100 mg) and cells were incubated for 24 hours, at 37°C and 5% $CO_2$. Exposure to light was minimized for all following steps. The cells were transfected with 0.5 µg HCN2 construct/ well, three wells per construct, and incubated for 24 hours. The medium was then replaced with medium containing CellMask Orange Plasma Membrane Stain and the plate incubated at 37 ° C for 15 min, followed by fixing for 1 hour at 37°C in 4% Paraformaldehyde. The cells were washed thrice with PBS, stained with DAPI (1:2000 dilution) and washed again thrice with PBS. Coverslips with the stained cells were mounted on slides in *SlowFade* Gold Antifade Mountant solution, sealed with clear nail varnish and stored in the dark at 4°C for 18 hours before imaging on a Zeiss LSM900. The images were collected at 20x overview and then at high resolution (magnification objective: 63x) using Airy scan. The imager was "blind" to the genotype of the constructs.

**Supplementary Table 1 Genomic findings of *HCN2* variants.**

| Genomic coordinates (GRCh37/hg19) | HGVS cDNA NM_031304.5 | HGVS Protein | Exon numbering | GnomAD Population Allele Frequencies (NFE/Total) & (Homozygote?) | Computational Prediction Scores | | | | Zygosity | ACMG Classification |
|---|---|---|---|---|---|---|---|---|---|---|
| | | | | | CADD | PROVEAN | Mutation Assessor | DANN | | |
| Ch19:590560C>A | c.615C>A | p.(His205Gln) | 1/8 | Not present | 24.6 | Damaging (-6,02) | Medium (3,225) | 0.9879 | Cpd het | Likely Pathogenic (PM2, PM3, PP3, PP4) |
| Ch19:603882G>A | c.971G>A | p.(Arg324His) | 2/8 | Not present | 25.8 | Damaging (-4,34) | Medium (3,42) | 0.9993 | Het | Pathogenic (PS3, PM2,PP3 PM5) |
| Chr19:603881C>T | c.970C>T | p.(Arg324Cys) | 2/8 | Not present | 26.1 | Damaging (-6.94) | Pathogenic (3.42) | 0.999 | Het | Likely Pathogenic (PS2, PM2, PM5, PP3) |
| Ch19:605092C>T | c.1088C>T | p.(Ala363Val) | 3/8 | Not present | 23.1 | Damaging (-3,36) | Medium (2,34) | 0.9989 | Het | Likely Pathogenic |

| | | | | | | | | | | |
|---|---|---|---|---|---|---|---|---|---|---|
| | | | | | | | | | | (PS2, PS3, PM1, PM2, PP3) |
| Chr19:605110A>G | c.1106A>G | p.(Asn369Ser) | 3/8 | Not present | 22.5 | Damaging (-4.37) | Low (1,905) | 0.9957 | Het | Likely Pathogenic (PS2, PM1, PM2, PP3) |
| Ch19:605124A>C | c.1120A>C | p.(Met374Leu) | 3/8 | Not present | 23.4 | Damaging (-2,62) | Low (1,635) | 0.957 | Het | Pathogenic (PS2, PS3, PM1, PM2, PP3) |
| Ch19:605134T>A | c.1130T>A | p.(Leu377His) | 3/8 | Not present | 27.0 | Damaging (-5,99) | Medium (3,395) | 0.9845 | Hom | Pathogenic (PS3, PM1, PM2, PP3) |
| Ch19:607971C>T | c.1226C>T | p.(Ser409Leu) | 4/8 | One occurrence in NFE | 23.3 | Damaging (-4,75) | Medium (2,19) | 0.9989 | Cpd het | Likely Pathogenic (PM2, PM3, PP3, PP4) |
| Ch19:608124G>A | c.1379G>A | p.(Gly460Asp) | 4/8 | Not present | NA | Damaging (-6,71) | Pathogenic (3,675) | 0,995 | Het | Pathogenic |

| | | | | | | | | | | |
|---|---|---|---|---|---|---|---|---|---|---|
| | | | | | | | | | | (PS2, PM1, PM2, PP3) |
| Ch19:608175delAGG | c.1432_1434del | p.(Glu478del) | 4/8 | Not present | NA | NA | NA | NA | Het | Likely Pathogenic (PS2, PM1, PM2, PP3) |
| Ch19:610299C>T | c.1478C>T | p.(Pro493Leu) | 5/8 | Not present | NA | Damaging (-9,7) | Medium (3,005) | 0.9984 | Hom | Likely Pathogenic (PS3, PM2, PP3) |
| Chr19:613423G>A | c.1760G>A | p.(Gly587Asp) | 6/8 | One occurrence in NFE | 27.0 | Damaging (-6.78) | Pathogenic (4.685) | 0.9986 | Hom | Pathogenic (PS3, PM2, PP3) |
| Ch19:613963insT | c.1936_1937insT | p.(Met647HisfsTer32) | 7/8 | Not present | NA | NA | NA | NA | Hom | Likely Pathogenic (PVS1, PM2, PM3) |

Abbreviations: ACMG, American college of medical genetics; Cmp Het, Compound Heterozygous; Het, Heterozygous; HGVS, human genome variation society; Hom, Homozygous; gnomAD, genome aggregation database; NFE, non-Finnish European; NA, not available.

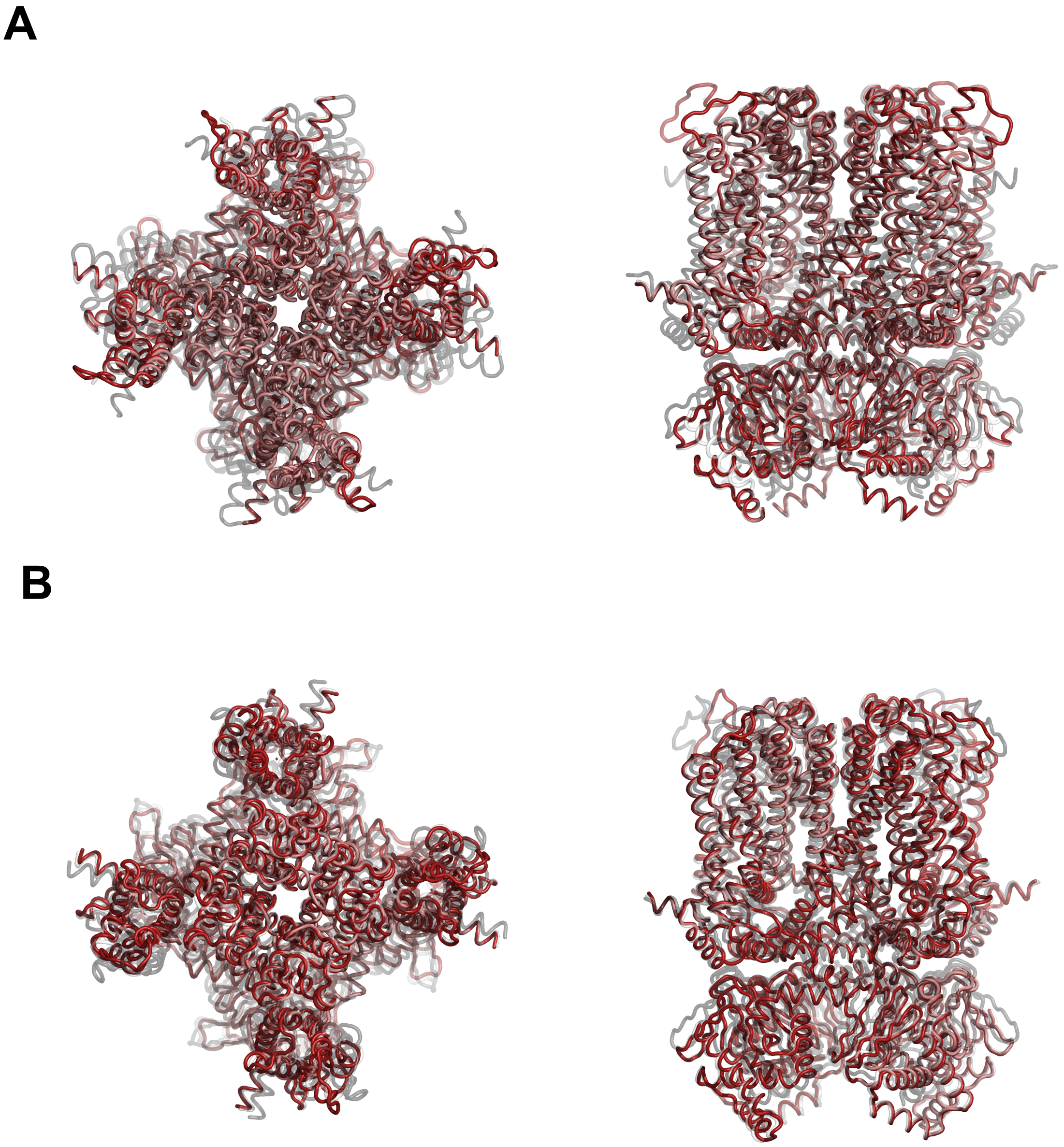


**Supplementary Figure 1 3D models of HCN2.** Molecular models of HCN2-wt in depolarized and hyperpolarized states were built from position 163 by homology with HCN1 (PDB 5U6P and 6UQF for resting and hyperpolarized models, respectively) with MODELLER V9.17. The alignment of the three structure was prepared with PyMOL (Molecular Graphics System, version 1.8, Schrödinger, LLC). The figure shows superimposed 3D-models of HCN1 (white), HCN2 (red) and HCN4 (grey, rabbit HCN4 (PDB 7NMN and PDB 6GYN)) as ribbons in both depolarized **(A)** and hyperpolarized states **(B)**. Left panels, top views; right panels, profile view.

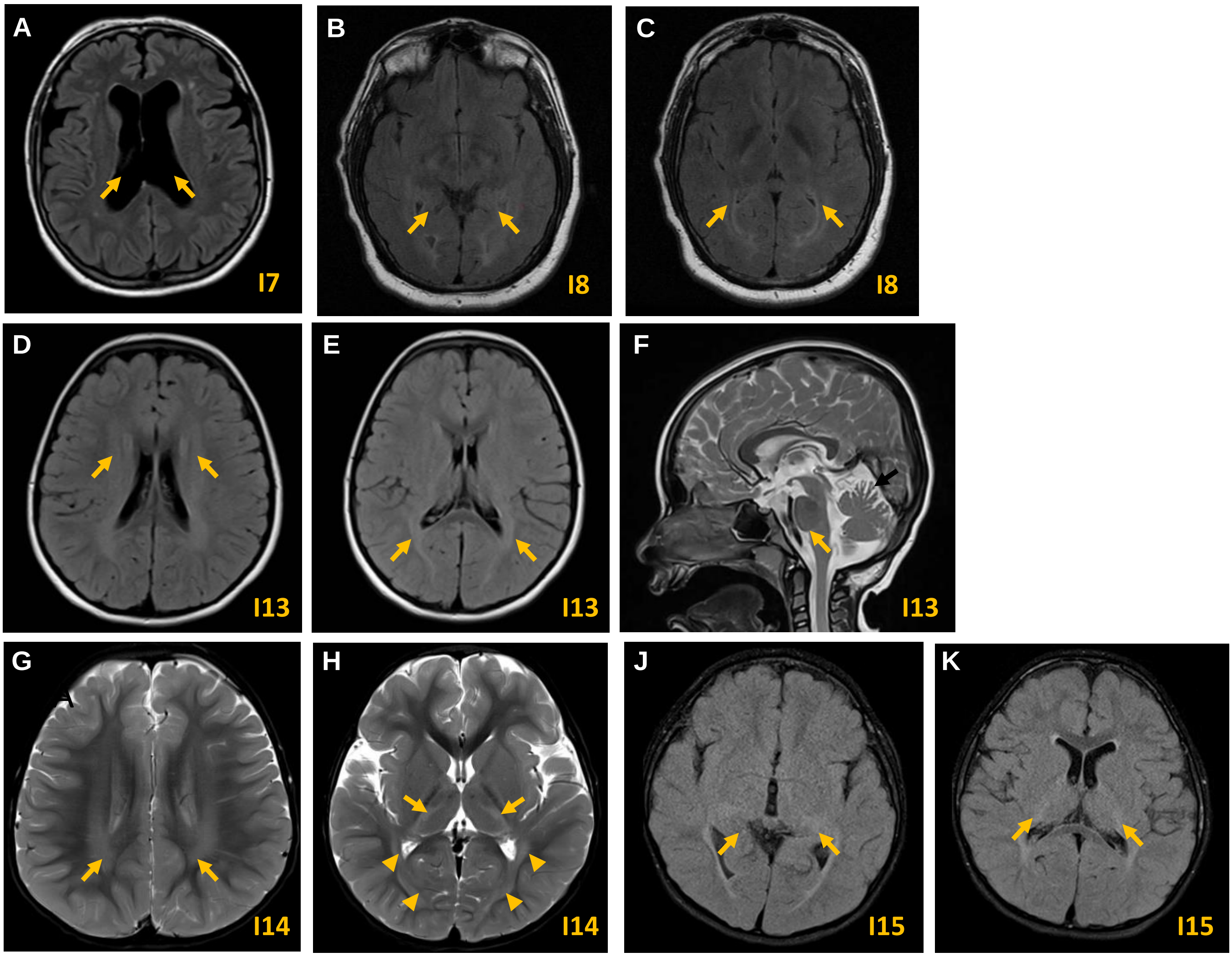


**Supplementary Figure 2 Cerebral MRI from individuals carrying p.(Ala363Val), p.(Asn369Ser) and p.(Glu478del) variants. (A)** Axial FLAIR MRI image from I7 show atrophy and increase of ventricular spaces (arrows). I7 carries p.(Ala363Val) variant. **(B,C)** Axial brain MRI images show mild T2 signal changes in the periventricular white matter posteriorly in I8, carrying p.(Asn369Ser) variant. **(D-H)** Axial MRI scans of individuals carrying the p.(Glu478del). **(D-F)** Axial FLAIR MRI images of I13 show periventricular white matter abnormalities. **(F)** Sagittal T2-weight image shows superior vermis atrophy (arrow C). **(G,H)** Axial T2-weighted image through the centrum semiovale of I14 demonstrates hyperintense signal involving the periventricular white matter with posterior predominance (arrows D). T2-weighted signal hyperintensity is also present in the ventrolateral thalami (E arrows), as well as the optic radiations and subcortical occipital white matter (E arrow heads). **(J,K)** Axial T2-weight images demonstrate posterior mild diffuse periventricular hyperintensity signal abnormality that extended into the posterior capsule into the thalami bilaterally in I15.

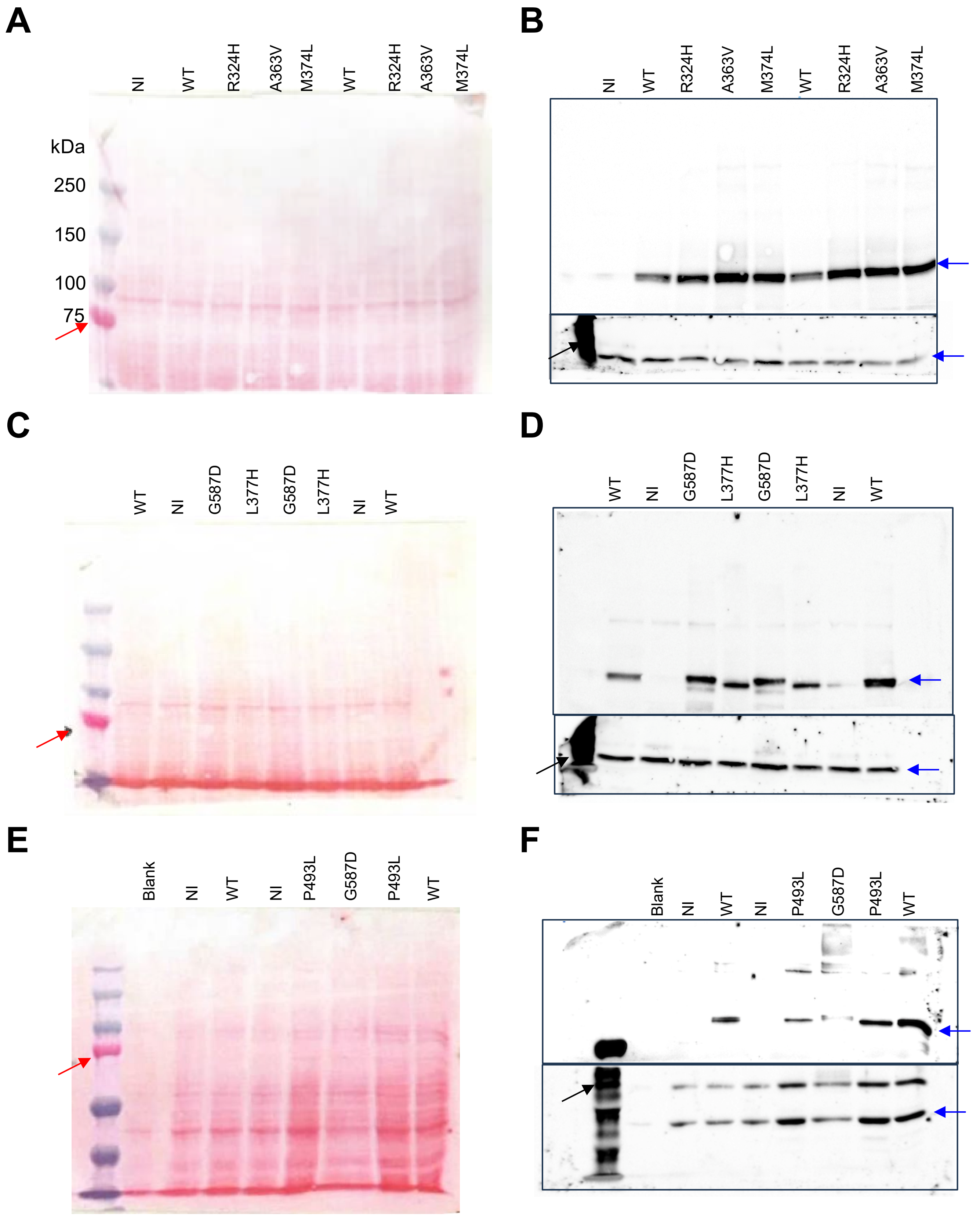


**Supplementary Figure 3 Western blot analysis of wt-HCN2 and its variants identified in this study.** Thirty µg of protein extracts from oocytes expressing wt-HCN2 and its variants were analyzed by immunoblotting. **(A,C,E)** Ponceau stained blot of protein extracts from non-injected oocytes (NI), wt-HCN2 controls (WT) and HCN2 variants (R324H, A363V, M374L, L377H, P493L and G587D). Red arrow indicates the position blot was cut prior to addition of primary antibodies. **(B,D,F)** Immunostained blots with anti-HCN2 (upper box) and anti-β-actin antibodies (lower box). Blue arrows shows cross-reacting protein with ~100 kDa for HCN2 and 42 kDa for β-actin. The slightly smaller L377H and P493L protein is assumed to result from a post-translational difference as no molecular reason could be detected by sequencing. The dark mark on the left is due to cross-reactivity with the size markers (black arrows).

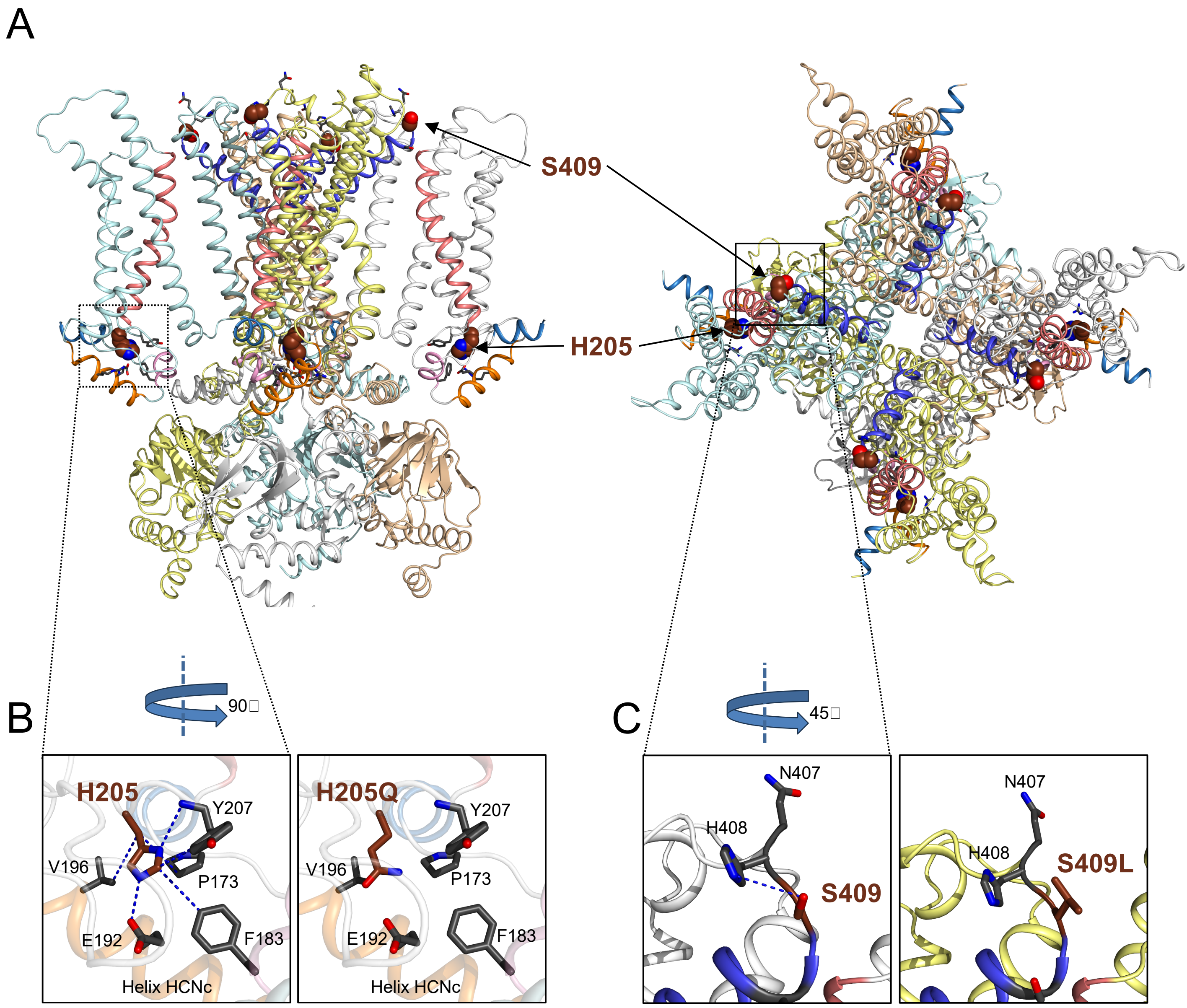


**Supplementary Figure 4 Structural analysis of p.(His205Gln)/p.(Ser409Leu) trans variant. A)** Top (left) and profile (right) views of HCN2 3D structure in depolarized state. His205 (spheres) is located at the N-terminal part of S1 and its side-chain is buried in a pocket formed by Pro173 (helix HCNa in light blue), Phe183 (helix HCNb in pink), Glu192 and Val196 (helix HCNc in orange) and Tyr207 (S1 in salmon). The side-chain of Ser409 faces the extracellular side close to Ser411. (**B,C**) The 3D structure of p.(His205Gln)/p.(Ser409Leu) trans variant was modelized V9.17 with MODELLER. **B)** Detail view of His205 environment in depolarized HCN2-wt (profile view). The side-chain of His205 (left panel) is highly stabilized by three interaction types : 1) polar interactions, a salt bridge with Glu192 and a hydrogen bond with Tyr207, 2) aromatic interactions with Phe183 and Tyr207, and 3) hydrophobic interactions with Pro173 and Ala196. The p.(His205Gln) variant (right panel) leads to the neutralization of a buried charge (His205), the disruption of 1) polar interactions with Glu192 and Tyr207, and 2) aromatic interactions with Phe183 and Tyr207. **C)** In depolarized state, the side-chain of S409 makes a hydrogen bond with His408, which is lost in p.(Ser409Leu) variant. Altogether, p.(His205Gln)/p.(Ser409Leu) trans HCN2 variant impact HCN2 structure stability.

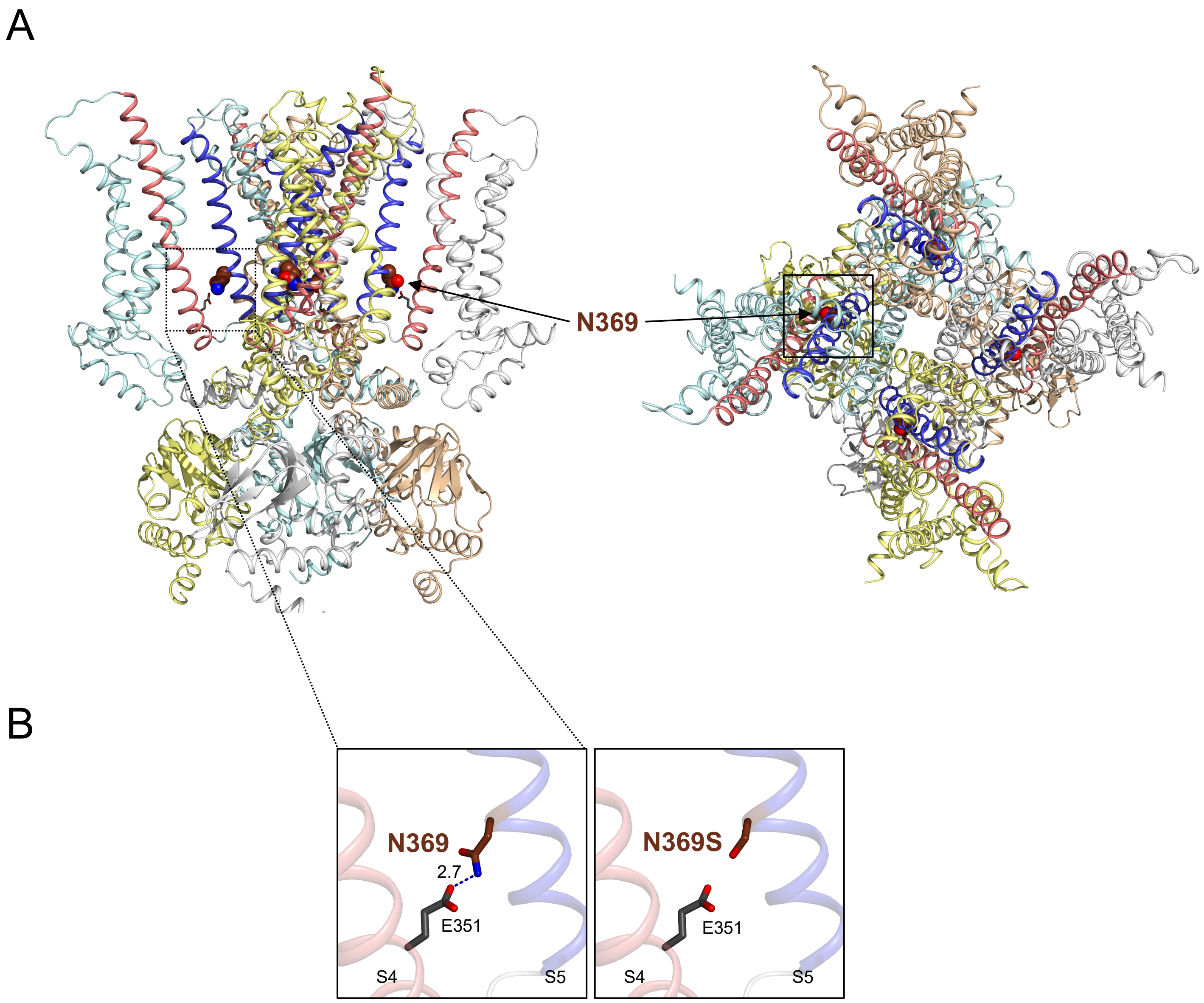


**Supplementary Figure 5 Structural analysis of p.(Asn369Ser) pathogenic variant. A)** Top (left) and profile (right) views of HCN2 3D structure in depolarized state. Asn369 (spheres) is located at the N-terminal part of S5 (in blue), and its side-chain faces S4 (in pink). (**B,C**) The 3D structure of p.(Asn369Ser) monoallelic variant was modelized V9.17 with MODELLER. **B)** Detail view of Asn369 environment in depolarized HCN2-wt (profile view). The side-chain of Asn369 (left panel) a hydrogen bond with Glu351. The p.(Asn369Ser) variant (right panel) leads to the disruption of the interaction with Glu351, and thus destabilizes the interface S4-S6. The distances are in Å.

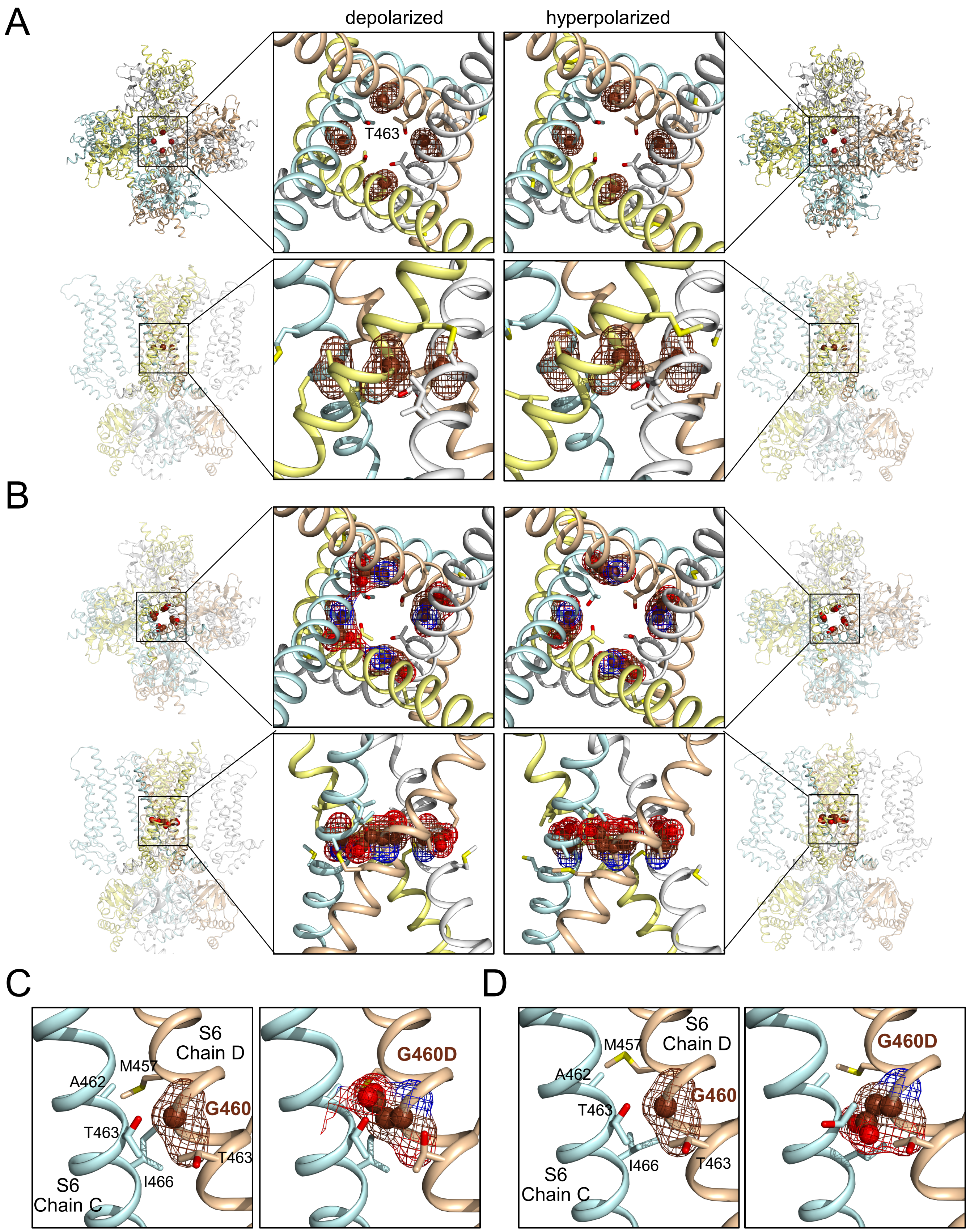


**Supplementary Figure 6 Structural analysis of p.(Gly460Asp) HCN2 pathogenic variant. (A,B)** From top to bottom, top and profile views of wt-HCN2 (**A**) and p.(Gly460Asp) variant (**B**) structures. G460 and G460D are represented in spheres with their electron density maps. In top views, the side chains of T463, close to G460 and G460D are facing the pore. G460 is located in S6 at the narrowest part of the pore. The p.(Gly460Asp) variant introduces a negative-charged and bulky residue, replacing a very small and uncharged residue, between two S6. (**C,D**) Profil views of the interface between two S6, showing G460 and G460D surrounded by M457, A462, T463 and I466, at depolarized **(C)** and hyperpolarized **(D)** states. For clarity, chain A and B were removed. The p.(Gly460Asp) variant structure shows clash between the Asp and M457, A462, T463 and I466. Thus, the p.(Gly460Asp) variant induces structural damage in HCN2.
